# Lifestyles and their relative contribution to biological aging across multiple organ systems: change analysis from the China Multi-Ethnic Cohort Study

**DOI:** 10.1101/2024.06.03.24308368

**Authors:** Yuan Zhang, Dan Tang, Ning Zhang, Yi Xiang, Yifan Hu, Wen Qian, Yangji Baima, Xianbin Ding, Ziyun Wang, Jianzhong Yin, Xiong Xiao, Xing Zhao

**Affiliations:** West China School of Public Health and West China Fourth Hospital, Sichuan University, Chengdu, China; Xiamen Center for Disease Control and Prevention, Xiamen, China; Chengdu Center for Disease Control and Prevention, Chengdu, China; School of Medicine, Tibet University, Lhasa, China; Chongqing Municipal Centre for Disease Control and Prevention, Chongqing, China; School of Public Health, the Key Laboratory of Environmental Pollution Monitoring and Disease Control, Ministry of Education, Guizhou Medical University, Guiyang, China; School of Public Health, Kunming Medical University, Kunming, China

**Keywords:** healthy lifestyle, biological aging, organ-specific aging, change analysis, Klemera-Doubal method of biological age

## Abstract

**Background:** Biological aging exhibits heterogeneity across multi organ systems. However, it remains unclear how is lifestyle associated with overall and organ-specific aging and which factors contribute most in Southwest China.

**Objective:** To examine the associations of healthy lifestyle with comprehensive and organ-specific biological ages and which lifestyle factors contribute most.

**Methods:** This study involved 8,396 participants who completed two surveys from the China Multi-Ethnic Cohort (CMEC) Study. The healthy lifestyle index (HLI) was developed using five lifestyle factors: smoking, alcohol, diet, exercise, and sleep. The comprehensive and organ-specific biological ages (BAs) were calculated using the Klemera-Doubal method based on longitudinal clinical laboratory measurements, and validation were conducted to select BA reflecting related diseases. Fixed effects model was used to examine associations between HLI or its components and the acceleration of validated BAs. We further evaluated the relative contribution of lifestyle components to comprehension and organ systems BAs using quantile G-computation.

**Results:** About two-thirds of participants changed HLI scores between surveys. After validation, three organ-specific BAs (the cardiopulmonary, metabolic, and liver BAs) were identified as reflective of specific diseases and included in further analyses with the comprehensive BA. The health alterations in healthy lifestyle index showed a protective association with the acceleration of all biological ages, with a mean shift of –0.19 (95%CI: –0.34, –0.03) in the comprehensive biological age acceleration. Diet and smoking were the major contributors to overall negative associations of five lifestyle factors with the comprehensive BA and metabolic BA accounting for 24% and 55% respectively.

**Conclusions:** Healthy lifestyle changes were inversely related to comprehensive and organ-specific biological aging in Southwest China, with diet and smoking contributing most to comprehensive and metabolic BA separately. Our findings highlight the potential of lifestyle interventions to decelerate aging and identify intervention targets to limit organ-specific aging in less-developed regions.

## Introduction

Aging is a global issue, with about 10 percent of the world population aged 65 years and over in 2022, and the proportion expected to be near 16 percent by 2050(1). In China, the proportion has reached 14.2% at the end of 2021(2). Aging leads to reduced life expectancy and chronic diseases through dynamic and heterogeneous changes in biological systems(3). While chronological age (CA) is widely used as a marker of aging, biological age (BA) has been constructed as a more accurate indicator for biological aging(4). Aging exhibits variations across and within individuals; people with the same CA can age at different rates, and distinct organ systems within an individual can also age differently(5). Comprehensive aging indicators like epigenetic clocks and PhenoAge, capturing different aging aspects, have been developed(6, 7). Recent studies have focused on organ-specific aging markers to understand their aging rate variations(8, 9). The process of biological aging, influenced by genetics, environment, and health behaviors, is modifiable, yet research has predominantly concentrated on comprehensive biological age (10–13). Nevertheless, current research on determinants of multi-organ biological aging remains limited. Further investigation is crucial to deepen understanding of comprehensive and organ-specific biological aging and to develop targeted interventions for age-related diseases of each organ system.

Lifestyle factors such as smoking, drinking, diet, physical activity, and sleep are recognized as contributors to many age-related diseases(14–16), and researchers have begun to explore how lifestyle factors influence the process of biological aging. Studies on lifestyle and BA can be summarized into two categories, one focusing on the association of a single lifestyle factor with biological aging(17–20) because lifestyle factors may be interrelated and have cumulative effects, the other further focusing on the impact of a combined lifestyle on biological aging. Research focusing on healthy lifestyles has been conducted in Chinese(21, 22), American(23), and British populations(24) and has found that adherence to a healthy lifestyle is consistently associated with slower biological aging. However, most studies used one-time measurements of lifestyle factors as exposure and could not examine the impact of lifestyle changes on BA. In addition, research has often focused on epigenetic age and PhenoAge. The Klemera-Doubal method of biological age (KDM-BA) has been shown to perform well in predicting frailty and mortality and is used to identify the biological ages of organ systems(8, 25). Therefore, KDM-BA, based on clinical-lab data sets, offers accessible and cost-effective means for aging identification and intervention. Besides, no study has been conducted to analyze further the relative contributions of multiple lifestyle changes on various organ systems BA, which could identify important influences on the comprehensive and organ-specific biological aging to prioritize interventions, especially in developing regions with limited resources. Southwest China is characterized by multi-ethnicity, unbalanced internal development, and rapid aging, making identifying priority intervenable aging factors important.

Therefore, based on the China Multi-Ethnic Cohort Study (CMEC), this study aimed to develop potential BAs for organ systems using the KDM algorithm, explore how lifestyle changes correlate with both comprehensive and organ-specific BAs, and further investigate the relative contributions of individual lifestyle components in southwest China.

## Methods

### Study Population

This study used data from the CMEC baseline and repeated surveys for analysis. The CMEC adopted a multi-stage stratified cluster sampling, with participants covering five provinces and seven ethnic groups (Tibetan, Yi, Miao, Bai, Bouyei, Dong, and Han) in Southwest China, representing the local regional and ethnic characteristics. The baseline survey of the CMEC was conducted between May 2018 and September 2019, and the repeated survey was conducted between August 2020 and June 2021. The CMEC baseline survey included 99,556 individuals, with 10% of the sample population selected for the repeated survey. Details of the CMEC procedures and methodology have been described in our previous study(26). Participants’ personal interviews, physical examinations, and clinical laboratory measurements were completed using a uniform standardized operating procedure in both surveys. The study protocol was approved by the Sichuan University Medical Ethical Review Board [ID: K2016038, K2020022]. All participants in the study signed an informed consent form before the investigation.

In this study, we included participants with available information on lifestyle, measurements for calculating BA required, and covariates. We excluded individuals with missing data to calculate BA and individuals with incomplete data on lifestyle and covariates for participants at the baseline and repeated survey. Finally, 8396 individuals aged 30-79 were included as the full set in the longitudinal analysis after matching (Supplementary Figure 1).

### Definition of lifestyle factors

Considering previous studies(15, 27), the healthy lifestyle index (HLI) was established through five lifestyle factors: smoking, alcohol consumption, diet, exercise, and sleep. Data for the lifestyle factors were obtained from the questionnaire, and the simplified semi-quantitative food frequency questionnaire (FFQ) was assessed by trained staff. The FFQ’s reproducibility and validity were evaluated by conducting repeated FFQs and 24-hour dietary recalls(28).

The definitions of a healthy lifestyle are as follows. Smoking was categorized according to smoking status, and never smoking was defined as healthy. For alcohol consumption, according to the findings of recent research on alcohol consumption in Chinese populations, being healthy was defined as being a non-regular drinker (drinking frequency less than once a week)(29). For diet, since the Mediterranean diet (MED) is currently the most internationally recognized healthy dietary pattern and has widely and robustly beneficial effects(30), we used the alternative Mediterranean diet (aMED) to evaluate dietary quality, with details shown in the supplemental material. Since the HLI already contained a drinking component, we removed the drinking item in the aMED, which had a score range of 7-35. We defined individuals with aMED scores ≥ population median as healthy diets. More details on the calculation of aMED can be found in previous studies(28). For exercise, based on how often participants participated in physical activity in their leisure time during the past year, regular exercise (“1–2 times/week”, “3–5 times/week,” or “daily or almost every day”) was categorized as healthy(31). For sleep, a sleep duration of 7-8 hours was defined as healthy sleep based on previous studies(32).

In addition, to comprehensively evaluate the impact of overall healthy lifestyle level on outcomes, we further constructed the HLI in this study. For each lifestyle, we assign 1 point to health and 0 points to the opposite. The HLI was calculated by directly adding up the five lifestyle scores, ranging from 0-5, with a higher score representing an overall healthier lifestyle, denoted as HLI (range) in the following text. We then transformed HLI into a dichotomous variable in this study, denoted as HLI (category), where a score of 4-5 for HLI was considered a healthy lifestyle, and a score of 0-3 was considered an unfavorable lifestyle that could be improved.

### Covariate Assessment

The selection of covariates is based on prior literature review and previously constructed directed acyclic graphs (Supplementary Figure 2). Covariates included age, sex, ethnicity (majority, minority), urbanicity (rural, urban), education (No schooling, primary school, Middle/high school, College/university), occupation (primary industry practitioner, secondary industry practitioner, tertiary industry practitioner, unemployed), marital status (married/cohabiting, not married/cohabitating), total energy intake (kcal/day), depression symptoms, anxiety symptoms, menopausal status in women (premenopausal, perimenopausal, postmenopausal), beverage intake (never, former consumer, currently consuming), dietary supplement intake, self-reported doctor-diagnosed diseases of diabetes, cardiovascular disease (CVD), and cancer. Among these, sex, ethnicity, urbanicity, and education were considered time-invariant variables, while other variables were considered time-varying.

### BA Construction

The comprehensive BA and BAs across multiple organ systems were estimated using the Klemera-Doubal method(33), which is well-validated in the Chinese population(34). We selected indicators for constructing BA from clinical-lab data sets measured in the baseline and repeated surveys of the CMEC, filtering based on a missing rate of less than 30%. Based on previous studies, these indicators were categorized into five systems based on the organ/system function they represent: cardiopulmonary, metabolic, liver, renal, and immune systems, as detailed in the Supplementary Table 2(8, 9). Thus, we developed a comprehensive BA alongside five organ system-specific BAs: cardiopulmonary BA, metabolic BA, liver BA, renal BA, and immune BA. The selection process was then completed following the methodology used in our previous studies, which are detailed in the supplementary materials.

After finishing the screening process, 15 measures were used in constructing the comprehensive BA, which were systolic blood pressure (SBP), waist-to-hip ratio (WHR), peak expiratory flow (PEF), γ-Glutamyl transpeptidase (GGT), albumin (ALB), low-density lipoprotein cholesterol (LDL-CH), high-density lipoprotein cholesterol (HDL-CH), triglyceride (TG), aspartate aminotransferase (AST), creatinine (Cr), alkaline phosphatase (ALP), urea, mean corpuscular volume (MCV), glycosylated hemoglobin (HBA1C) and platelet count (PLT). The cardiopulmonary BA was assessed using SBP and PEF; the metabolic BA was assessed using LDL-CH, HDL-CH, HBA1C, TG and WHR; the liver BA was assessed using AST, GGT, ALP, ALB; the renal BA was assessed using Cr and urea, and the immune BA was assessed using PLT and MCV. Using the above measures, we calculated each BA in the male and female populations separately using the KDM algorithm. To quantify variation in biological aging between participants, we calculated BA acceleration, which is the difference between each BA and CA simultaneously.

### BA Validation

To assess the performance of the newly developed BAs of multiple organ systems, we performed a validation analysis to evaluate the relationships between them and organ-specific diseases. Given the short follow-up period in the CMEC study, we conducted a cross-sectional analysis using baseline data. We separately examined the associations between the comprehension BA and CVD, diabetes, cancer, cardiopulmonary BA and CVD and chronic bronchitis, the metabolic BA and CVD and diabetes, the liver BA and chronic hepatitis or cirrhosis, the immune BA and rheumatoid arthritis. For interpretability, we simultaneously standardized and classified each BA acceleration into categories (|BA acceleration| ≤ 1, BA acceleration < –1, and BA acceleration > 1). We used logistic regression to assess the associations between each continuous and categorized BA (|BA acceleration| ≤ 1 serving as the reference group) and the diseases. Since the values of BA acceleration of the cardiopulmonary BA were small, it was not processed as categorical variables. The model was additionally adjusted for five healthy lifestyle factors based on the covariates. The comprehensive BA constructed in the CMEC study has been validated to better reflect age-related disease and frailty(35). After completing the validation analysis, we selected BAs reflecting aging and related diseases for further analysis.

### Statistical analysis

We described changes in lifestyle between the two waves of surveys, where changes were categorized as becoming healthier, remaining unchanged, and becoming unhealthier. For single lifestyles, remaining unchanged indicates that the factor has remained healthy or unhealthy at two surveys; becoming healthier suggests that it was unhealthy at baseline and became healthy on the repeated survey, and vice versa. For the HLI scores, no change indicates the same score in both surveys, a change to healthier implies a repeated survey score higher than the baseline score, and a shift in less healthy indicates a repeated survey score lower than the baseline score. The dichotomous HLI’s change categorization was similar to the single lifestyle. The baseline characteristics of participants were described across categories in the dichotomized HLI, with continuous variables represented by median (Interquartile range, IQR) and categorical variables by proportions (Table 1).

**Table 1.**
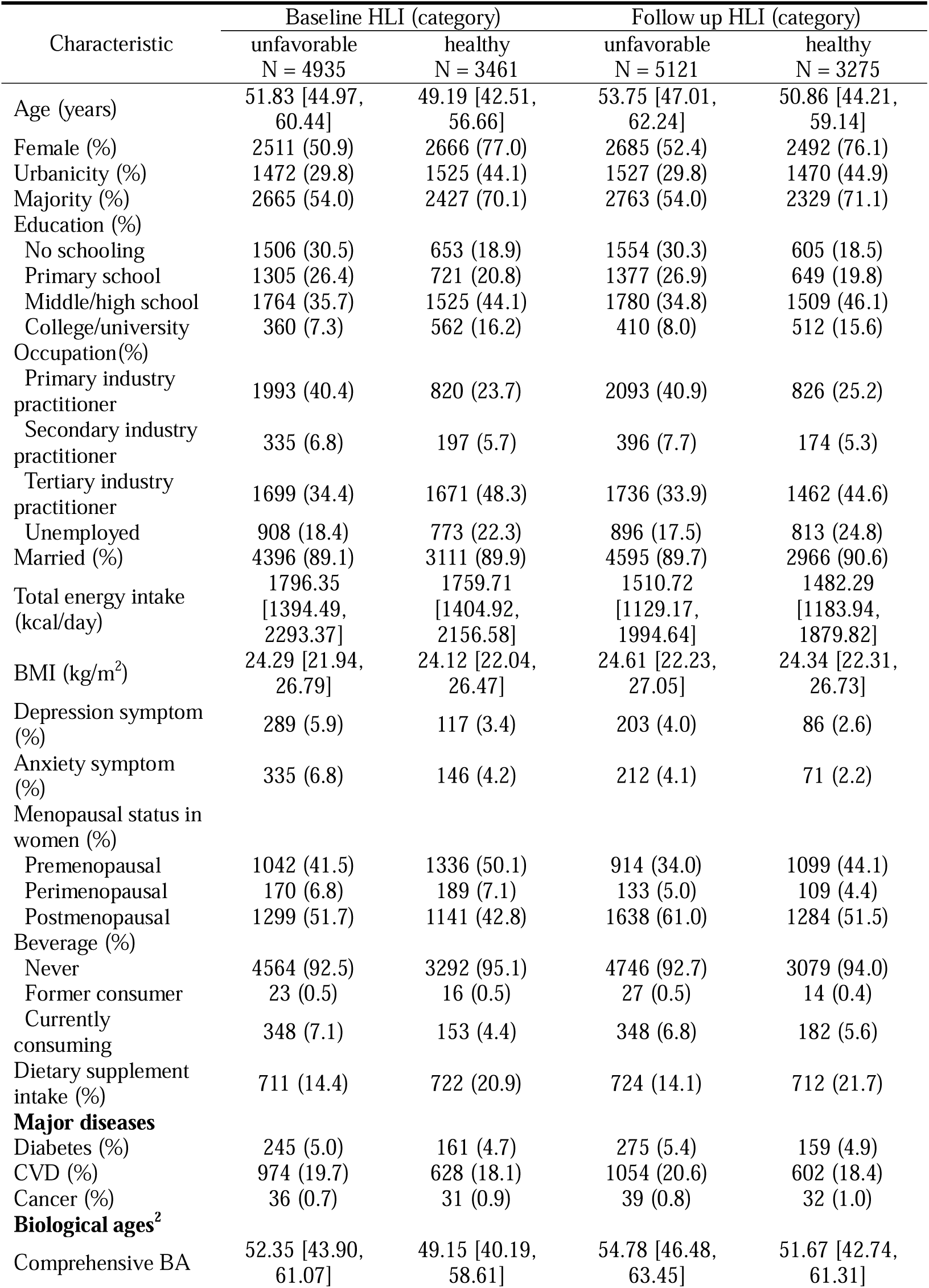
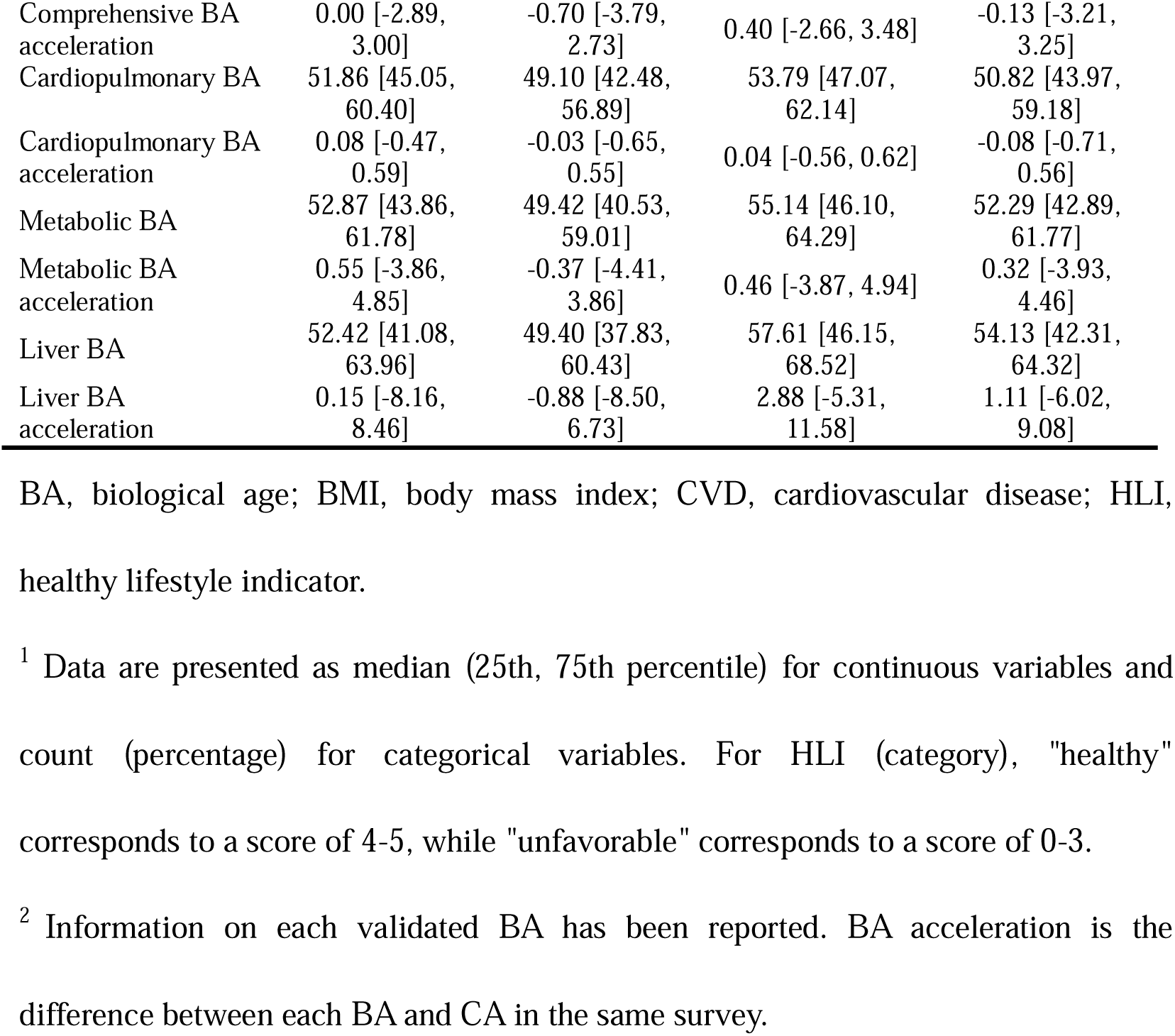
Characteristics of participants in baseline and repeated surveys, based on the categorized HLI^1^.

We used the fixed effects model (FEM) to examine associations between each lifestyle factor and the acceleration of validated BAs for people who participated in both surveys. FEM is extensively applied in the analysis of panel data in the fields of sociology, economics, and public health research, where it serves to control for unobserved individual heterogeneity. The detailed methodology of the FEM is outlined in the supplementary materials. Because of the approximately 2-year interval between the two CMEC waves, we assumed that the short-term effect of lifestyle changes on biological aging was linear during this period. Multivariable-adjusted models were constructed to account for potential confounding, including time-varying and time-invariant variables and baseline CA. We incorporated five lifestyle factors as exposures simultaneously into the model, while the continuous and binary HLI were entered into the model as exposures separately. Since the BAs we analyzed may potentially measure overlapping aspects of human aging, we did not correct for multiple comparisons (Supplementary Figure 3).

To further obtain the relative contribution of each lifestyle factor on validated comprehension and organ systems BA, the present study used data from two waves for quantile G-computation (QGC) analysis. QGC is a statistical technique for evaluating the effects of mixture exposures, capable of discerning positive and negative influences, and is widely used in environmental epidemiology research(36, 37). Additionally, subgroup analyses for continuous and categorized HLI and each validated BA were conducted across sex (male vs. female), baseline age (<60 vs.≥60), ethnicity (majority vs. minority), urbanicity (rural vs. urban), baseline BA acceleration (<0 vs. ≥0), and baseline disease status (free of diabetes, CVD, and cancer vs. either one). The heterogeneity between strata was assessed using the Q test (α = 0.1 was considered significant heterogeneity).

We performed sensitivity analyses concerning exposure definitions, confounders, and our analysis method. First, we repeated the analysis of the association between lifestyles and BAs using a standard FEM, with adjustments made only for time-varying variables. Second, we used alternative common health criteria for each lifestyle factor separately, and we repeated the FEM analyses and QGC analyses by replacing the health definition of one lifestyle factor at a time. Exposure definitions for the main analyses and sensitivity analyses are shown in the Supplementary Table 1. Third, we did not adjust for body mass index (BMI) in models because some studies have suggested that BMI may be an intermediate factor between lifestyles and health outcomes. However, in the sensitivity analysis, we additionally adjusted for BMI in the FEM and QGC analyses models. Statistical analyses were performed using R Project for Statistical Computing version 4.1.1 (Vienna, Austria).

## Results

### Lifestyle changes between two waves in CMEC

Figure 1 shows the changes in lifestyle factors in the study population from baseline to repeated surveys. For the HLI (range), more than 60 percent of participants experienced a lifestyle change. For the HLI (category), about 30 percent of participants changed lifestyle categories, and the percentages were close for both types. For each lifestyle factor, the proportion of individuals who remained unchanged during the two surveys was higher than that of individuals who changed, with smoking and drinking having the highest proportion of unchanged individuals at more than 90 percent, indicating that these two lifestyle factors are less likely to change. Sleep and diet behaviors had a relatively high percentage of change, accounting for over one-third, and exercise had a slightly lower rate of change, nearly 30 percent.

**Figure 1.**
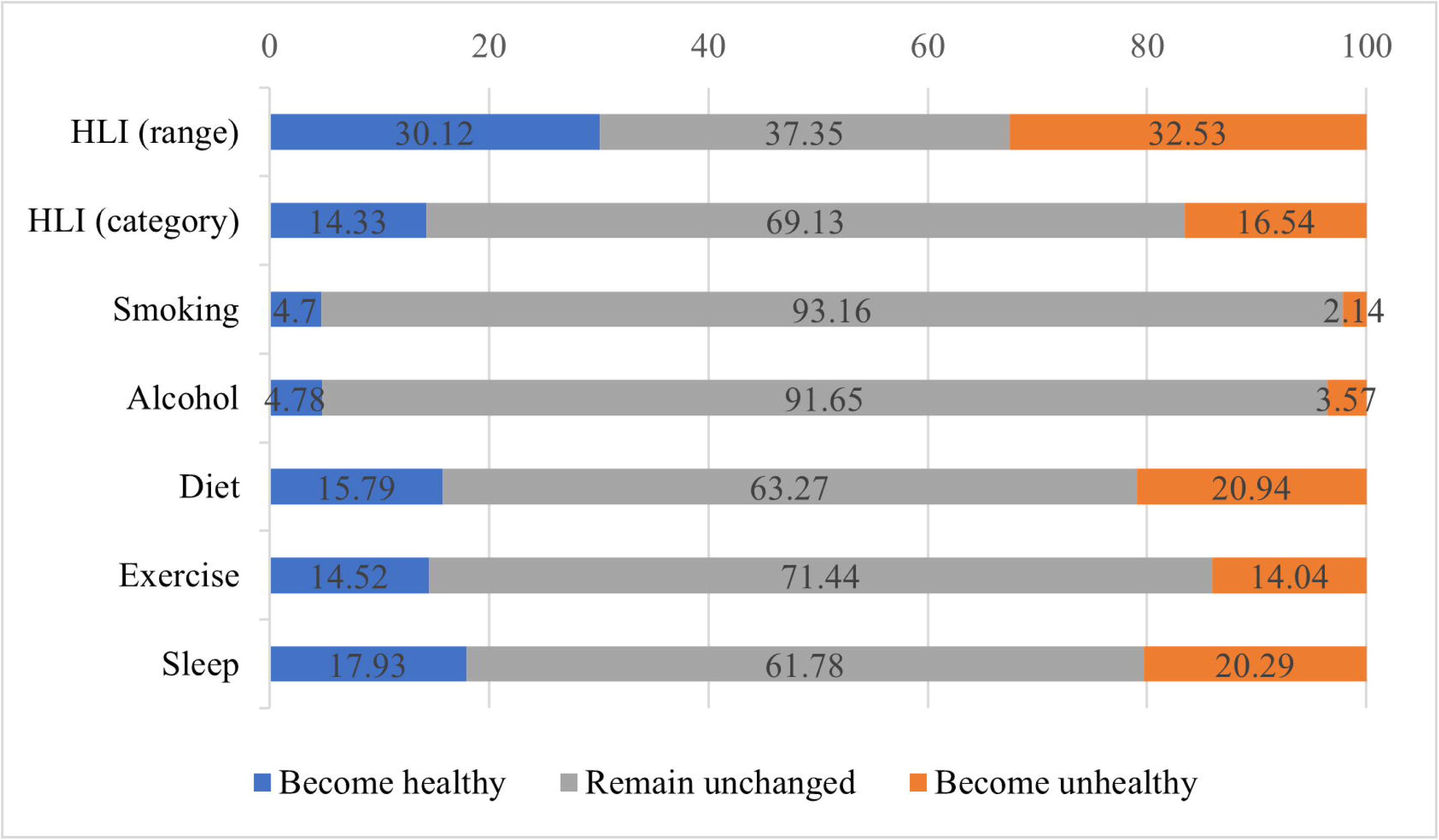
Changes in lifestyle factors from baseline to repeated survey. HLI, healthy lifestyle indicator. The blue bars represent the percentage of individuals who have transitioned to healthier behaviors or improved their HLI score; the gray bars represent the percentage of individuals whose behaviors or HLI score have not changed; the orange bars represent the percentage of individuals who have transitioned to unhealthier behaviors or whose HLI score has decreased.

### Construction and validation of organ systems BAs

Details of the comprehensive and organ systems BAs are presented in Supplementary Table 3. The results of the cross-sectional analysis between the comprehensive and organ systems BAs and diseases are presented in Supplementary Table 4. Accelerated comprehensive BA was positively correlated with an increased risk of CVD and diabetes. The cardiopulmonary BA, the liver BA, and the metabolic BA were each linked to an increased risk of their respective diseases. An increase in cardiopulmonary BA acceleration was associated with a higher risk of cardiovascular diseases but not with chronic obstructive pulmonary disease. Per 1-SD increase of the liver BA was associated with a higher risk of chronic hepatitis or cirrhosis (OR 1.28, 95% C/ 1.23-1.33), and per 1-SD increase in metabolic BA corresponded to a higher risk of CVD (OR 1.21, 95% C/ 1.19-1.23) and diabetes (OR 3.23, 95% C/ 3.12-3.34). However, no expected associations with related diseases were observed for immune BA. Finally, we concluded that the cardiopulmonary BA, the metabolic BA, and the liver BA could reflect organ-specific disease and were included in subsequent analyses along with comprehensive BA.

### Characteristics of participants by HLI categories

Table 1 reports the characteristics of populations categorized by dichotomous HLI at baseline and the repeated survey. In this study, 3461 individuals had a healthy lifestyle, and individuals with an unfavorable lifestyle accounted for a higher proportion (n (%) = 4935 (58.8%)) at baseline. There was a slight decrease in the proportion of people with a healthy lifestyle at the time of the repeated survey (n (%) = 3275 (39.0%)), while 5049 (61.0%) had an unfavorable lifestyle.

Since the lifestyle categories of participants may change, the distribution of population characteristics between baseline and repeated surveys changes slightly but is generally similar. Participants with healthy lifestyles were more likely to be female, in urban areas, Han Chinese, with higher levels of education, less likely to be employed in primary and secondary industries, more likely to be premenopausal and with dietary supplement intake, less likely to have anxiety, depression, and younger in biological age, as shown by the lower median of BA and BA acceleration of the comprehensive, cardiopulmonary, metabolic and liver BA. For major diseases, the proportion of people with a healthier lifestyle was closer to that of people with an unfavorable lifestyle in both surveys.

### Associations of lifestyle factors and HLI changes with BAs acceleration and relative contributions

Table 2 shows the associations of individual lifestyle factors and continuous and dichotomous HLI with all validated BAs acceleration. For comprehensive BA, a significant negative association was found between HLI and accelerated biological aging, with a mean change of –0.19 (95% C/: –0.34, –0.03) in the BA acceleration for a healthier lifestyle change. Results showed that a change of –0.15 (95% C/: –0.29, – 0.00) in the comprehensive BA acceleration was associated with a shift in diet. For the cardiopulmonary BA, metabolic BA, and liver BA, the HLI was associated with a reduction in each BA acceleration, although some of the results were not statistically significant. Shift in smoking was related to a change in metabolic BA acceleration of – 0.54 (95%C/: –0.97, –0.11), and the results of individual lifestyle factors and the cardiopulmonary BA as well as the liver BA did not reach statistical significance.

**Table 2.** Associations of healthy lifestyle factors and HLI with the BA acceleration of validated BAs.

| Variables | Comprehensive<br>BA acceleration<br>$\beta$ (95%CI) | Cardiopulmonary<br>BA acceleration<br>$\beta$ (95%CI) | Metabolic<br>BA acceleration<br>$\beta$ (95%CI) | Liver<br>BA acceleration<br>$\beta$ (95%CI) |
| --- | --- | --- | --- | --- |
| HLI (range) |  |  |  |  |
| Per 1-point increase | -0.11 (-0.19, -0.03) | -0.02 (-0.04, -0.01) | -0.12 (-0.22, -0.02) | -0.23 (-0.47, 0.00) |
| HLI (category) |  |  |  |  |
| To have a healthy lifestyle | -0.19 (-0.34, -0.03) | -0.03 (-0.07, 0.00) | -0.16 (-0.36, 0.03) | -0.19 (-0.66, 0.27) |
| Healthy lifestyle factors |  |  |  |  |
| ΔSmoking | -0.13 (-0.47, 0.21) | -0.03 (-0.10, 0.04) | -0.54 (-0.97, -0.11) | -0.57 (-1.58, 0.44) |
| ΔAlcohol | -0.17 (-0.47, 0.13) | -0.02 (-0.09, 0.04) | -0.18 (-0.56, 0.21) | -0.59 (-1.50, 0.31) |
| ΔDiet | -0.15 (-0.29, -0.00) | -0.01 (-0.04, 0.02) | -0.18 (-0.36, 0.00) | -0.10 (-0.53, 0.33) |
| ΔExercise | -0.16 (-0.32, 0.00) | -0.03 (-0.07, 0.00) | -0.09 (-0.30, 0.11) | -0.05 (-0.53, 0.44) |
| ΔSleep | -0.02 (-0.16, 0.12) | -0.02 (-0.05, 0.01) | 0.01 (-0.17, 0.19) | -0.36 (-0.78, 0.06) |

BA, biological age; HLI, healthy lifestyle indicator.
ΔDiet, change in dietary quality between the baseline and repeated survey; ΔExercise,
change in exercise between the baseline and repeated survey; ΔSleep, change in sleep
between the baseline and repeated survey; CI, confidence interval.
Estimates were obtained using FEMs treating the BA accelerations as the dependent
variables and HLI (as either continuous or as categorized) or five individual lifestyle
factors as the independent variables. Models were adjusted for age, occupation,
marital status, total energy intake, depression symptoms, anxiety symptoms,
menopausal status in women, beverage intake, dietary supplement intake, diabetes,
cardiovascular disease, cancer, sex, ethnicity, urbanicity, education, and the

The relative contribution of each lifestyle factor on the comprehensive and the cardiopulmonary BA, the metabolic BA, and the liver BA acceleration based on the QGC method are shown in Figure 2. For comprehensive BA, the relative contribution of diet was 0.24. Although the relative contributions of drinking and exercise were slightly higher than that of diet, they were not statistically significant. For metabolic BA, smoking was the major contributor, weighting 0.55. For cardiopulmonary BA and liver BA, exercise and alcohol consumption were the most contributing components in all lifestyle factors separately, although the results were statistically significant.

**Figure 2.**
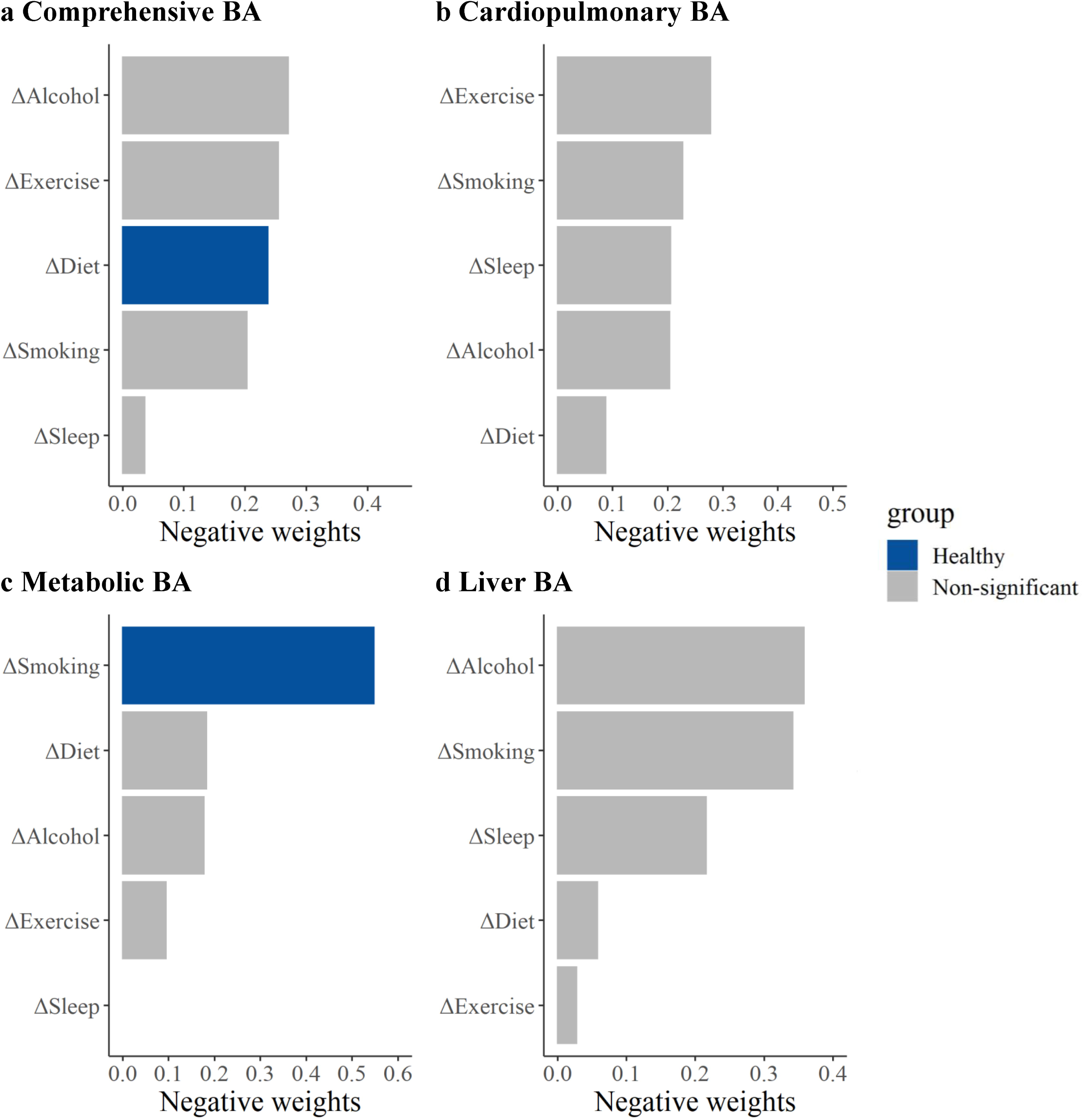
Relative contributions of five healthy lifestyle components to BAs acceleration. Panels: Results of the comprehensive BA acceleration, the cardiopulmonary BA acceleration, the metabolic BA acceleration, and the liver BA acceleration (A–D). ΔSmoking, change in smoking status between the baseline and repeated survey; ΔAlcohol, change in alcohol consumption between the baseline and repeated survey; ΔDiet, change in dietary quality between the baseline and repeated survey; ΔExercise, change in exercise between the baseline and repeated survey; ΔSleep, change in sleep between the baseline and repeated survey. Estimates were obtained using QGC, which treated the BA accelerations as the dependent variables and five individual lifestyle factors as the independent variables. Models were adjusted for age, occupation, marital status, total energy intake, depression symptoms, anxiety symptoms, menopausal status in women, beverage intake, dietary supplement intake, diabetes, cardiovascular disease, cancer, sex, ethnicity, urbanicity, education, and the participants’ age at baseline. The blue bars represent results that are statistically significant in the FEM analysis, while the gray bars represent results in the FEM analysis that were not found to be statistically significant and positive weights were not shown.

### Results of subgroup analysis and sensitivity analysis

Figure 3 shows the results of the subgroup analyses of the HLI range and category with the comprehensive BA acceleration. For the HLI range, the direction of the association was consistent with the whole population, with statistical significance only in females, the younger, minorities, rural residents, accelerated BA group, and no baseline disease populations. The negative association was stronger in minorities (Heterogeneity test *P* = 0.028). For categorized HLI and AA, the direction of the association was broadly consistent with the whole population. The continued HLI results for cardiopulmonary BA showed a stronger negative association in females (Heterogeneity test *P* = 0.045, Supplementary Figure 4). For metabolic BA and liver BA, no differences were observed between different subgroups (Supplementary Figure 5 and 6).

**Figure 3.**
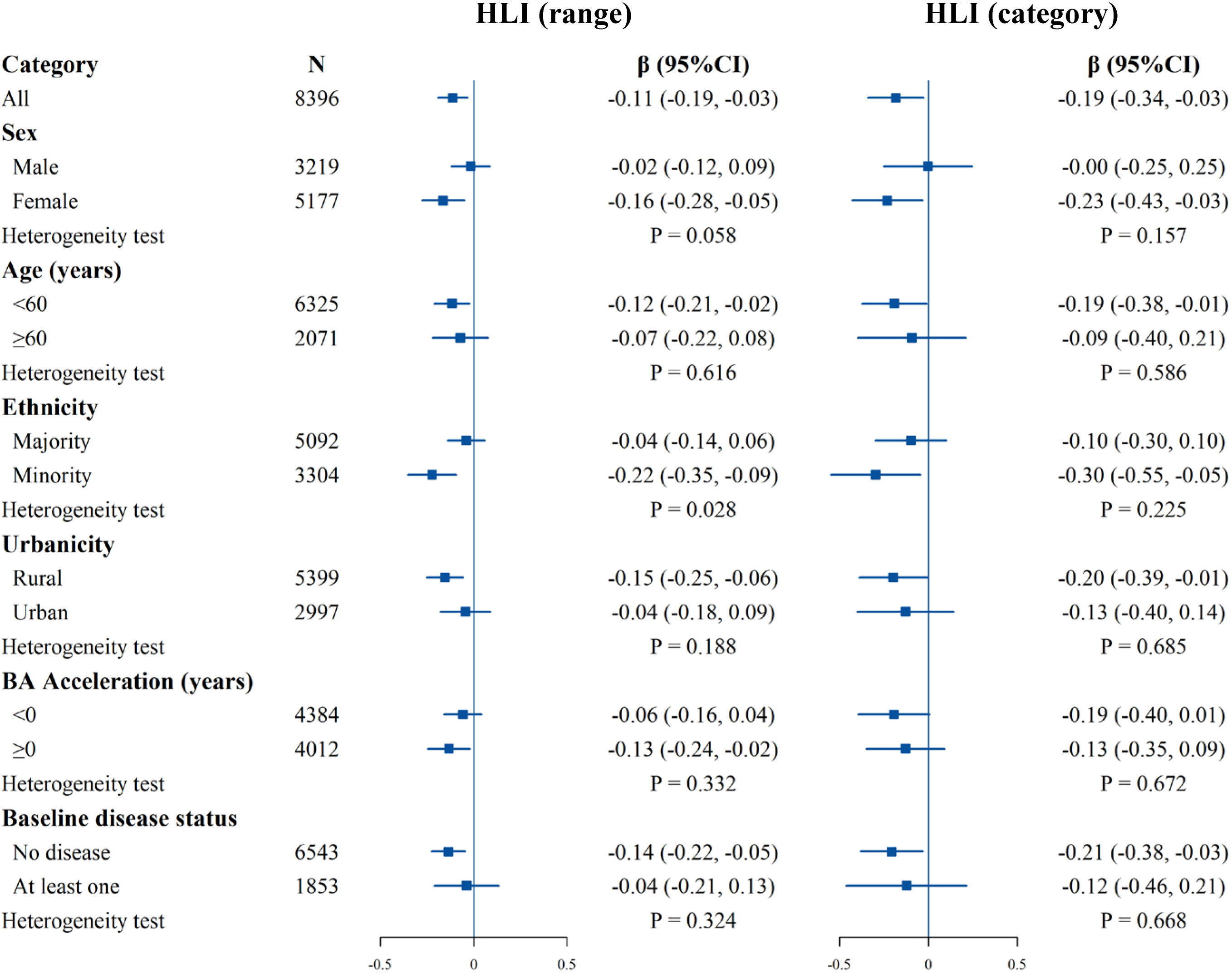
Stratified analysis of estimated associations between the HLI and the comprehensive BA acceleration. All models were adjusted for age, occupation, marital status, total energy intake, depression symptoms, anxiety symptoms, menopausal status in women, beverage intake, dietary supplement intake, diabetes, cardiovascular disease, cancer, sex, ethnicity, urbanicity, education, and the participants’ age at baseline, with exclusion of the stratified variable as appropriate. The boxes represent point estimations. Horizontal lines represent 95% CI.

We conducted a series of sensitivity analyses to test the robustness of the results. Firstly, the results of the standard FEM analysis were broadly consistent with the primary analysis (Supplementary Table 5 and Supplementary Figure 7). For the comprehensive BA, exercise along with diet was negatively correlated with BA acceleration, and in the main analysis, exercise was borderline statistically significant. While the results of some lifestyles for liver function BA changed, none were statistically significant. Secondly, changing the definition of health for any lifestyle factor had negative association estimates with all BAs acceleration for the HLI. Although the effect size of some lifestyle factors changed, most of the results remained stable. Full results are reported in Supplementary Table 6 and Supplementary Figure 8-11. Thirdly, with additional adjustment for BMI, the direction and magnitude of the associations were generally consistent with the main analysis results (Supplementary Table 7 and Supplementary Figure 12). In summary, the sensitivity analysis results aligned with the main results and were relatively robust.

## Discussion

This study examined the association of lifestyle changes on biological aging in the Southwest Cohort based on two surveys. About two-thirds of the participants were observed to change their HLI scores between the two surveys. The health alterations in HLI showed a protective association between accelerated aging of comprehension and each organ system. The relative contribution of diet was the largest among all lifestyle components for the comprehensive BA, and smoking contributed the most to the metabolic BA, respectively, suggesting that these factors are particularly crucial in specifically decelerating the pace of biological aging.

In our study, the prevalence of healthy lifestyles was higher than the prevalence reported by research based on the China Kadoorie Biobank study and the China Nutrition and Health Surveillance (38), with those with 4-5 lifestyle factors accounting for 41.2% and 39.0% at baseline and repeated surveys, respectively. The relatively high prevalence of healthy lifestyles in this study may be explained by he recent establishment of the CMEC and an overall trend towards healthier lifestyles, alongside different lifestyle standards. Furthermore, we noted lifestyle changes in our research and subsequently employed the FEM for analysis based on this observation. Lifestyle may be influenced by environmental factors such as urbanization and individual characteristics such as health awareness, and thus, lifestyle may change over time, yet fewer studies have focused on this(39). The high proportion of changes occurring in sleep, exercise, and diet here suggests that they may be more variable and relatively easy to intervene. For overall health styles, there was a higher proportion of changes in lifestyle scores but fewer shifts from unhealthy to healthy or healthy to unhealthy. That is, while lifestyle fluctuates over time, it is less likely to change substantially, consistent with the findings of another study(14, 40).

Our study reveals that healthy lifestyle shifts can slow the pace of comprehensive BA, with diet being a crucial component. Both the HLI range and category showed robust protective associations. Although there are differences in lifestyle definitions and aging indicators, available studies consistently find that healthier lifestyles are associated with lower aging acceleration(21, 23, 41, 42). Despite limited research on how lifestyle components contribute to aging, our study provides additional evidence. We used the widely recommended aMED as a healthy dietary criterion, which includes rich plant foods, olive oil, moderate fish, and red wine, some of which may slow aging and promote health(43). In our study, the association between exercise and comprehensive BA was nearly statistically significant. Higher levels of exercise have been shown to be associated with a lower incidence of chronic disease and a longer lifespan(44, 45). Diet and exercise may slow aging through beneficial DNA methylation changes(46), modification of gut microbiota(47), and reducing oxidative stress and inflammation(48, 49), collectively enhancing fitness and slowing the aging pace. Combining the previous randomized controlled trial with our findings, the evidence suggested that diet and exercise are easily modifiable anti-aging factors in the real world(13).

Another contribution of our study is presenting evidence of different lifestyle factors influencing multi-organ BA, which was constructed and validated for Southwest China. The HLI still played a protective role against aging across various organ systems, yet the components contributing most varied specifically. Biological aging varies across organ systems, and the associations with lifestyle may differ. Change in smoking played a significant role in metabolic aging. The current study of the British population has also identified smoking as a factor influencing aging across multiple organ systems(9). The Coronary Artery Risk Development in Young Adults Study revealed that smoking contributed to epigenetic aging acceleration by 83.5%(41). The variation in results could be attributed to the fact that current studies offer evidence of cumulative exposure to smoking(17, 21), while our research focuses on short-term changes. Short-term smoking behavior changes may be sufficient to impact metabolism aging but do not significantly affect the comprehensive BA. Existing research has shown that the metabolic and immune systems can impact various systems through inter-organ aging pathways, making it essential to focus on modifiable factors of metabolic system aging. Hence, we discovered that smoking could serve as an intervention target for organ system aging, holding the potential to delay the onset of diseases in specific systems, thereby extending a healthy lifespan.

In subgroup analysis, we discovered that negative associations of HLI and the comprehensive BA acceleration varied among ethnic groups. No observed differences in HLI category change between ethnic groups, potentially due to insufficient statistical power. Compared to the majority group, the minority may have lower socioeconomic status and less health awareness, leading to less management of lifestyle factors and, consequently, stronger associations when changes in these factors occur. Given that biological aging is a cause of various long-term outcomes, these results highlight the importance of conducting interventions among minority groups in Southwest China.

## Strengths and limitations

Unlike previous studies that used one measurement of lifestyle or biological age, our study utilized longitudinal data from two waves of the CMEC study, which measured the longitudinal changes in lifestyle behaviors and BA of the body and multi-organ systems. We also assessed the impact of alterations in healthy lifestyle factors on validated BAs and further characterized the relative contributions. This comprehensive approach offers the latest evidence for the early prevention and targeted behavioral interventions of organ-specific aging and related diseases in southwestern China.

This study has certain limitations. Firstly, our construction of multi-organ BA was limited by the clinical lab data measures, preventing the development of BAs for some organ systems like the brain. Additionally, we could not fully capture specific aging processes of the immune system because of few immune-related indicators and the possible omission of critical markers. However, we used routinely detected biomarkers to construct the BA, which are cost-effective and easily implemented. Therefore, this multi-organ BAs are suitable for broad adoption in primary prevention, especially in developing areas like Southwestern China. Besides, the CMEC had a limited range of diseases collected, precluding all BAs validation. Thus, we finally included only those BAs that could be validated for further analysis. Secondly, as an observational study, we established a prospective association between healthy lifestyles and accelerated aging, yet it does not confirm a causal relationship. Thirdly, the CMEC was established in 2018 with only data from two survey waves, which highlights the drawback of reduced statistical power of FEM. Future research could build on long-term, multi-wave follow-ups, employing FEM analysis based on repeatedly collected data to derive more reliable causal effect estimates. Fourthly, there is no gold standard for measuring biological aging. However, the KDM-BA method used in this study has the advantage of accurately predicting aging-related outcomes, and it has been validated in our study population(35). Fifth, assessment of lifestyle factors was based on self-reported data collected through questionnaires, which may be subject to recall bias. Lastly, our study is based on a population from the Southwestern China which could limit the generalizability of our findings. However, the Southwestern China is characterized by its diverse ethnicities and lower developmental level, with few studies addressing biological aging there. This research may provide crucial information for interventions in other less-developed regions.

## Conclusions

In summary, within the Southwest China population, we observed that healthy lifestyle changes were inversely related to comprehensive and organ-specific accelerated biological aging, with diet and smoking making the most significant contributions to the comprehensive BA and metabolic BA separately. These findings underscore the potential for lifestyle interventions to slow the aging pace and identify priority intervention targets to various organ-specific aging in less-developed regions.

## Acknowledgements

We sincerely thank all the participants and staff of the CMEC study. We gratefully acknowledge Professor Xiaosong Li, the former principal investigator of CMEC research, for his leadership and tremendous contribution to the establishment of CMEC. Professor Li passed away in 2019.

## Authors’ contributions

DT, JY and XX designed the study. YZ and DT analyzed the data and drafted the paper. All other authors were involved in the collection of the data. All authors made significant contributions to drafting and revising the manuscript and have approved the final version for publication.

## Data availability

Data described in the manuscript, code book, and analytic code will be made available by contacting the corresponding author.

## Ethics approval and consent to participate

This study was approved by the Sichuan University Medical Ethical Review Board [ID: K2016038, K2020022]. All procedures in the study were consistent with the 1964 Declaration of Helsinki and its subsequent amendments. All subjects agreeing to take part in the study signed informed written consent.

