## Supplementary methods, figures and tables for "Lifestyles and their relative contribution to biological aging across multiple organ systems: change analysis from the China Multi-Ethnic Cohort Study"

The construction process of the biological ag

Dietary assessment

Statistical methods

Fixed effect model

Quantile G-computation

Supplementary figures

Supplementary Figure 1. Flowchart of the study

Supplementary Figure 2. The final constructed directed acyclic graph (DAG)

Supplementary Figure 3. Heat map of correlations between the comprehensive and multiple organ systems BAs

Supplementary Figure 4. Stratified analysis of estimated associations between the HLI and the cardiopulmonary BA acceleration.

Supplementary Figure 5. Stratified analysis of estimated associations between the HLI and the metabolic BA acceleration.

Supplementary Figure 6. Stratified analysis of estimated associations between the HLI and the liver BA acceleration.

Supplementary Figure 7. Relative contributions of five healthy lifestyle components to BAs acceleration with adjustment for time-varying covariate

Supplementary Figure 8. The relative contributions of five healthy lifestyle components to the comprehensive BA acceleration with altered healthy lifestyle criteria

Supplementary Figure 9. The relative contributions of five healthy lifestyle components to the cardiopulmonary BA acceleration with altered healthy lifestyle criteria

Supplementary Figure 10. The relative contributions of five healthy lifestyle components to the metabolic BA acceleration with altered healthy lifestyle criteria

Supplementary Figure 11. The relative contributions of five healthy lifestyle components to the liver BA acceleration with altered healthy lifestyle criteria

Supplementary Figure 12. Relative contributions of five healthy lifestyle components to the comprehensive and the metabolic BA acceleration with additionally adjusted for BMI

Supplementary tables

Supplementary Table 1. Detailed definitions of lifestyle factors in the main analysis and sensitivity analysis

Supplementary Table 2. Candidate indicators used to construct the comprehensive and multi organ systems of biological age

Supplementary Table 3 Description of BA and BA acceleration

Supplementary Table 4. Associations of the comprehensive and multi organ systems BA acceleration with organ-specific diseases

Supplementary Table 5. Associations of healthy lifestyle factors and HLI with the BA accelerations with adjustment for time-varying covariates

Supplementary Table 6. Associations of healthy lifestyle factors and HLI with the BA accelerations altered healthy lifestyle criteria

Supplementary Table 7. Associations of healthy lifestyle factors and HLI with the BA accelerations with additionally adjusted for BMI

Supplementary reference

**Supplementary methods**

**The construction process of the biological age**

We constructed the comprehensive biological age (BA) and multi organ systems BA using the validated Klemera-Doubal method, based on clinical biomarkers and anthropometric measurements in the CMEC. We selected indicators for constructing BA measured in both the baseline and repeated surveys, filtering based on a missing rate of less than 30%. These indicators were categorized into five systems based on the organ/system function they represent: cardiopulmonary, metabolic, liver, renal, and immune systems, as detailed in Table S2. The construction of BAs was mainly divided into the following steps:

Step 1: Normality tests were conducted on the measurements of biomarkers. For those not meeting normality criteria, the Box-Cox transformation was applied to achieve normal distribution.

Step 2: To select biomarkers of aging, variables unrelated to participants' chronological age (CA) were excluded based on criteria of $p>0.05$ and $|r|<0.1$.

Step 3: Excluded redundant biomarkers that might reflect the same aspects of aging based on the existing knowledge and correlation among those biomarkers.

A correlation matrix was generated to examine the relationships between biomarkers and to eliminate potential redundant variables, especially those from the same organ or system with very high correlation coefficients ($r\geq0.70$).

Through the above steps, we selected 15 biomarkers, including systolic blood pressure (SBP), waist-to-hip ratio (WHR), peak expiratory flow (PEF), γ-Glutamyl transpeptidase (GGT), albumin (ALB), low-density lipoprotein cholesterol (LDL-CH), high-density lipoprotein cholesterol (HDL-CH), triglyceride (TG), aspartate aminotransferase (AST), creatinine (Cr), alkaline phosphatase (ALP), urea, mean corpuscular volume (MCV), glycosylated hemoglobin (HBA1C) and platelet count (PLT).

Step 4: Using the above measures, we calculated each BA in the male and female populations separately. This algorithm converts biomarkers to aging rates, making them comparable in two steps. The first step is to regress each biomarker against the CA. The second step is to summarize the age estimates for each biomarker and the CA to construct the BA. The formula is shown below:

$$\begin{aligned} {BA}_{EC}=\frac{\sum_{j=1}^{m} \left( x_{j}-q_{j} \right)\left( \frac{k_{j}}{s_{j}^{2}} \right)+\frac{CA}{s_{BA}^{2}}}{\sum_{j=1}^{m} \left( \frac{k_{j}}{s_{j}} \right)^{2}+\frac{1}{s_{BA}^{2}}}\# \end{aligned}$$

$x_{j}$ was the $j$th selected biomarker value, $m$ was the number of biomarkers. We constructed the regression of the $j$th biomarker in the reference population to chronological age (CA). And $k_{j}$, $q_{j}$ and $s_{j}$ represent the slope, intercept and root mean squared error, respectively. $s_{BA}^{2}$ was the estimated variance in chronological age explained by the selected biomarkers in the reference population. $\mathrm{BA}_{EC}$ was the estimated KDM-BA.

Thus, we developed a comprehensive BA alongside five organ system-specific BAs, namely comprehensive BA, cardiopulmonary BA, metabolic BA, liver BA liver, renal BA, and immune BA. To quantify variation in biological aging between participants, we calculated BA acceleration, which is the difference between each BA and CA at the same time.

**Dietary assessment**

We employed the alternative Mediterranean diet (aMED) and the plant-based diet index (PDI) as standards for a healthy diet in our study. Using adapted methodologies based on dietary data from CMEC, we assessed these two a priori dietary patterns for all participants. Participants' dietary intakes were recorded at baseline and follow-up using a validated, simplified semi-quantitative food frequency questionnaire (FFQ). This simplified FFQ had 13 comprehensive representative food groups used for collecting the consumption and frequency of subjects’ dietary intake during the year preceding the interview. Intake frequencies were offered in four options: daily, weekly, monthly, or annually. Reported average food intakes, provided in reference portion sizes or grams, were subsequently converted into weekly consumption figures in grams. We evaluated the FFQ's reproducibility and validity by conducting repeated FFQs and 24-hour dietary recalls. Intraclass correlation coefficients for reproducibility ranged from 0.15 for fresh vegetables to 0.67 for alcohol, while deattenuated Spearman rank correlations for validity ranged from 0.10 for soybean products to 0.66 for rice.

Our calculated aMED score incorporates eight components: vegetables, legumes, fruits, whole grains, fish, the ratio of monounsaturated fatty acids (MUFA) to saturated fatty acids (SFA), red and processed meats, and alcohol. Each component's consumption was divided into sex-specific quintiles. Scores ranging from 1 to 5 were assigned based on quintile rankings to each component, except for red and processed meats and alcohol, for which the scoring was inverted. The alcohol criteria for the aMED was defined as moderate consumption. Since the healthy lifestyle index (HLI) already contained a drinking component, we removed the drinking item in the aMED, which had a score range of 7-35 with a higher score reflecting better adherence to the overall Mediterranean dietary pattern. We defined individuals with aMED scores ≥ population median as healthy diets.

We calculated the PDI using 15 food groups, which were then categorized into plant foods and animal foods. The plant foods category includes tubers, fresh vegetables, soy products, fresh fruits, whole grains, tea, vegetable oil, pickled vegetables, and refined grains. The animal foods category consists of red and processed meats, poultry and products, seafood, eggs and products, dairy and products, and animal oil. Scores ranging from 1 to 5 were assigned based on quintile rankings to each plant foods, for animal foods the scoring was inverted. Therefore, a higher intake frequency of plant foods and a lower intake frequency of animal foods result in an increased PDI.

**Statistical methods**

**Fixed effect model**

Fixed effects models, also referred to as concurrent change-change analysis, are extensively applied in the analysis of panel data in the fields of sociology, economics, and public health research, where they serve to control for unobserved individual heterogeneity. It could eliminate those unmeasured time-invariant factors by analyzing changes and therefore reduce the bias from unmeasured time-invariant confounding. The formulas are as follows:

$$\begin{aligned} y_{i0}=\beta x_{i0}+\gamma z_{i0}+\boldsymbol{\alpha}_{\boldsymbol{i}}+u_{i0}\#\left( 1 \right) \end{aligned}$$

$$\begin{aligned} y_{i1}=\beta x_{i1}+\gamma z_{i1}+\boldsymbol{\alpha}_{\boldsymbol{i}}+u_{i1}\#\left( 2 \right) \end{aligned}$$

By subtracting equation (1) from equation (2):

$$\begin{aligned} \Delta y_{i} \sim\beta\Delta x_{i}+\gamma\Delta z_{i}\#\left( 3 \right) \end{aligned}$$

where $\boldsymbol{\alpha}_{\boldsymbol{i}}$ represent time-invariant confounders and they were eliminated through the differential.

In our study, we utilized this model with the change in the comprehensive BA and multi organ system BAs acceleration between baseline and follow-up as the dependent variable and the change of lifestyle factors between baseline and follow-up as the exposure variable. However, to enhance the flexibility of our models and account for potential variations in the effects of time-invariant variables and CA, as has been commonly done in previous studies, we additionally adjusted for time-invariant variables and baseline value of CA (see formula (4) below).

$$\begin{aligned} \Delta y_{i} \sim\beta\Delta x_{i}+\gamma\Delta z_{i}+\rho\alpha_{i}\#\left( 4 \right) \end{aligned}$$

$\Delta y_{i}$: change in KDM-BA acceleration between two surveys

$\Delta x_{i}$: change in tea consumption status between two surveys

$\alpha_{i}$: time-invariant covariates

$\Delta z_{i}$: change in time-variant covariates between two surveys

**Quantile G-computation**

We used quantile G-computation (QGC) from the qgcomp package in R to estimate the relative contributions of a single lifestyle factor to comprehension and organ systems BA. The QGC obtains causal relationships and estimates weights for each component, which has been widely used in epidemiological research. In our study, the QGC method was carried out through the following steps:

Step 1: Arrangement of component data. The component data could keep the original scale, or be converted into categorized coded data as required, such as in quartiles. For the convenience of explanation, we directly used scores (0 or 1) for each lifestyle factor in the current study.

Step 2: Fitting regression models. The required covariates could be included in the model, which was omitted here for the sake of concise symbolic expression. The model was as follows:

$$Y=\beta_{0}+\sum_{j=1}^{k} \beta_{j}X_{j}^{q}+\epsilon$$

In the current study, $k$ represented the total number of lifestyle components, and $\epsilon$ represented the residual term. $x_{j}^{q}$ was the score of $j$th lifestyle component. $\sum_{j}^{k} \beta_{j}$ was the mixture effect of the total lifestyles. Weight of component or relative contribution was defined as ${\beta_{j}}/{\sum_{j}^{k} \beta_{j}}$. It could be interpreted as the contribution of the $j$th component to the total effect when all the lifestyle components change from unhealthy to healthy simultaneously. Because the estimation process of the above model did not limit the positive or negative of $\beta_{j}$, we could estimate the positive or negative weight of each component at the same time. When there were both positive and negative associations between components and outcome, the positive and negative weights were calculated separately, with all positive weights summing to 1 and all negative weights summing to 1.

**Supplementary figures**

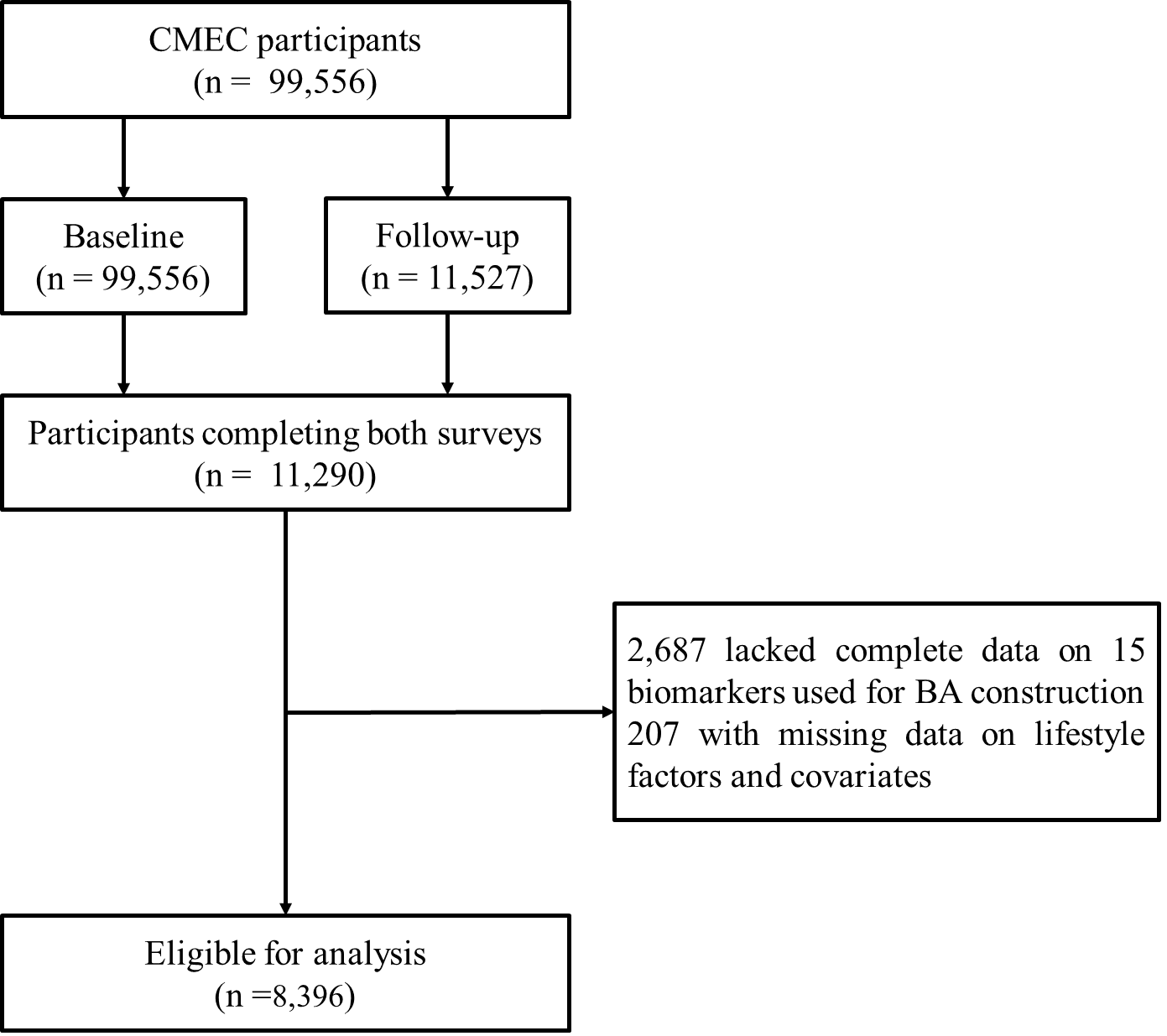

**Supplementary Figure 1. Flowchart of the study**

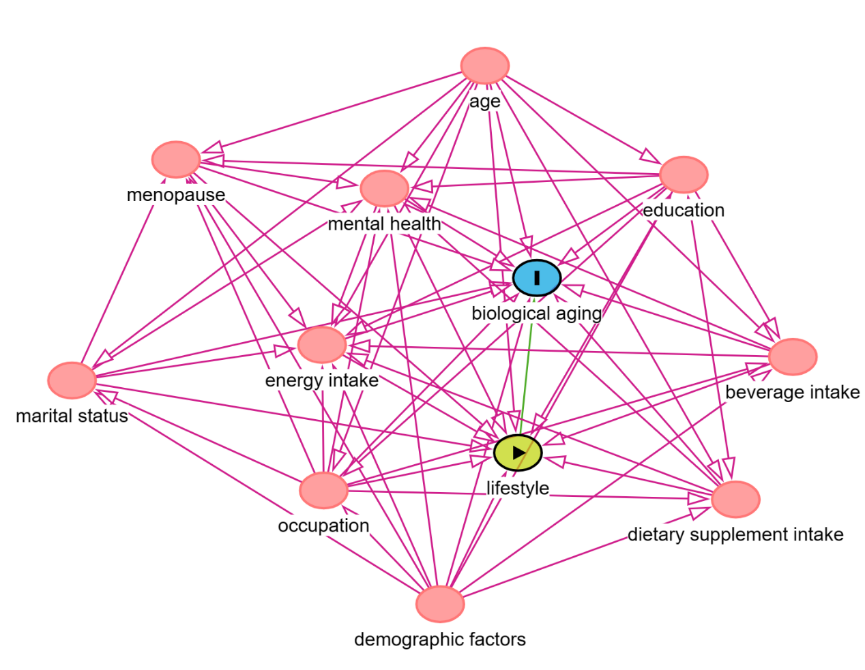

**Supplementary Figure 2. The final constructed directed acyclic graph (DAG)**

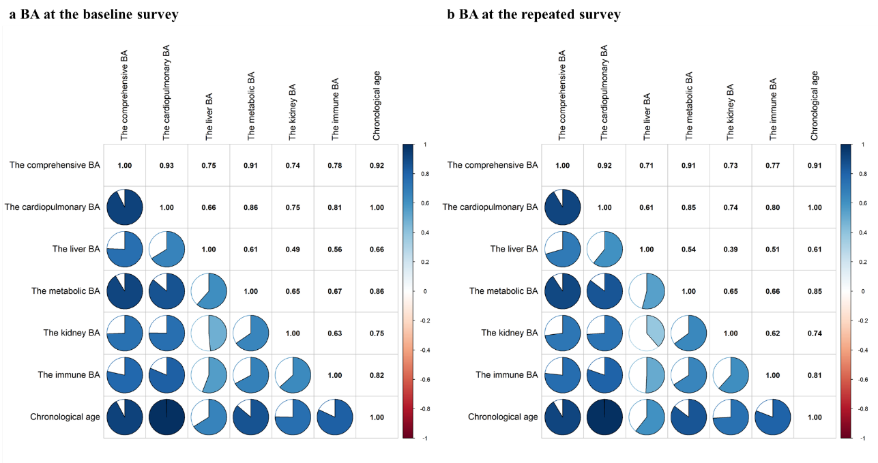

**Supplementary Figure 3. Heat map of correlations between the comprehensive and multiple organ systems BAs**

The color gradient displays Spearman correlation coefficients, with blue denoting positive correlations and red indicating negative correlations.

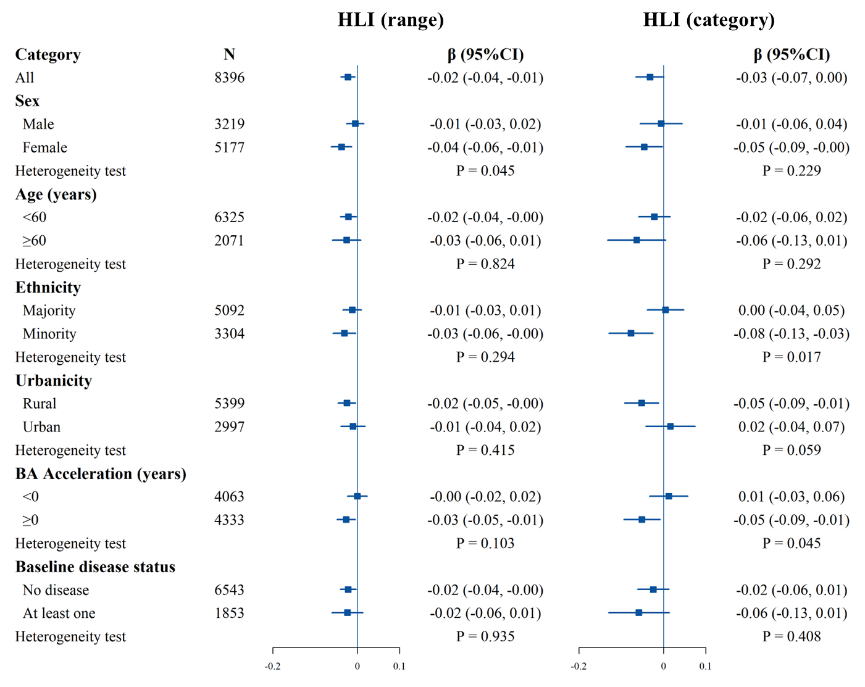

**Supplementary Figure 4. Stratified analysis of estimated associations between the HLI and the cardiopulmonary BA acceleration.**

All models were adjusted for age, occupation, marital status, total energy intake, depression symptoms, anxiety symptoms, menopausal status in women, beverage intake, dietary supplement intake, diabetes, cardiovascular disease, cancer, sex, ethnicity, urbanicity, education, and the participants' age at baseline, with the exclusion of the stratified variable as appropriate. The boxes represent point estimations. Horizontal lines represent 95% CI.

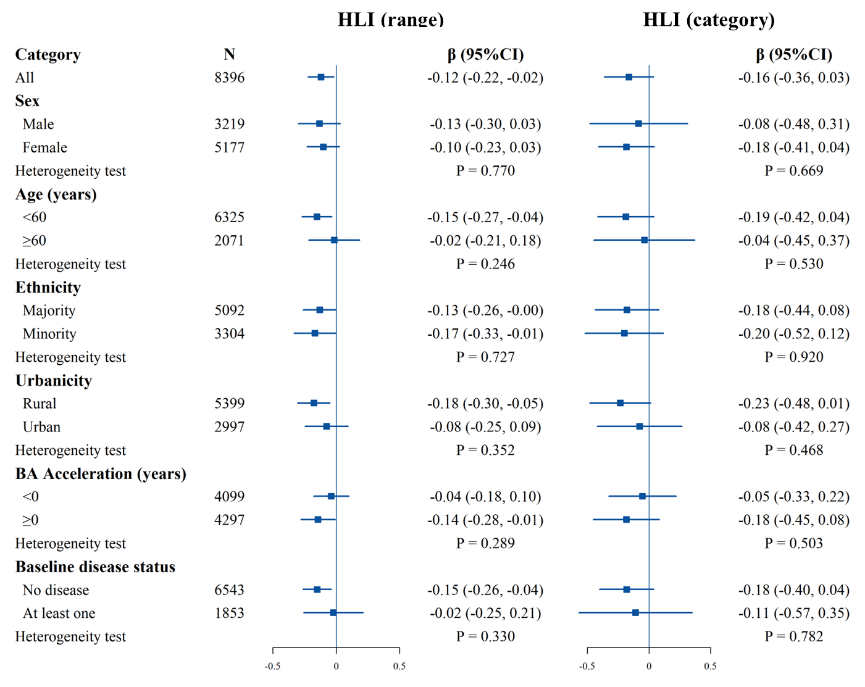

**Supplementary Figure 5. Stratified analysis of estimated associations between the HLI and the metabolic BA acceleration.**

All models were adjusted for age, occupation, marital status, total energy intake, depression symptoms, anxiety symptoms, menopausal status in women, beverage intake, dietary supplement intake, diabetes, cardiovascular disease, cancer, sex, ethnicity, urbanicity, education, and the participants' age at baseline, with the exclusion of the stratified variable as appropriate. The boxes represent point estimations. Horizontal lines represent 95% CI.

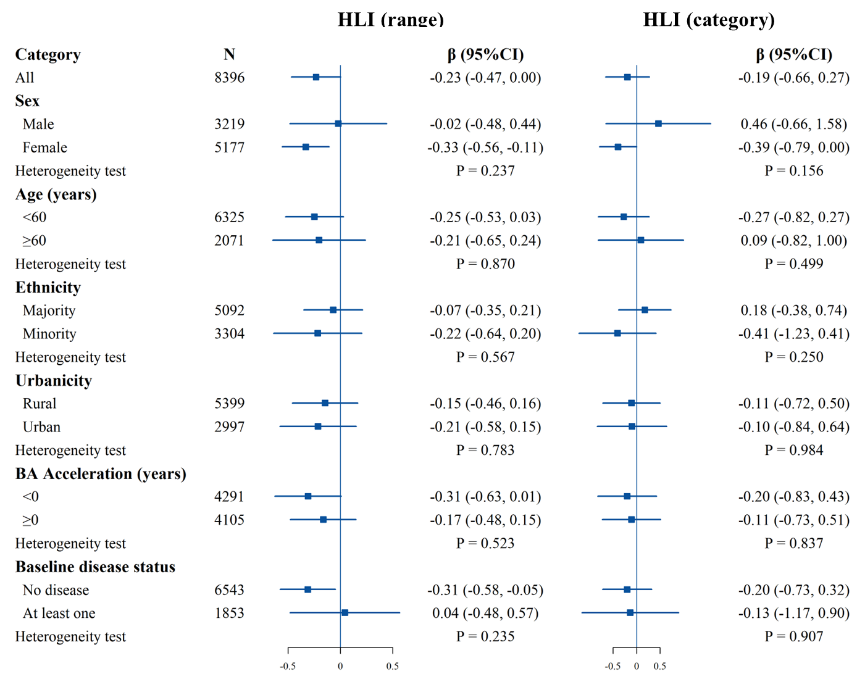

**Supplementary Figure 6. Stratified analysis of estimated associations between the HLI and the liver BA acceleration.**

All models were adjusted for age, occupation, marital status, total energy intake, depression symptoms, anxiety symptoms, menopausal status in women, beverage intake, dietary supplement intake, diabetes, cardiovascular disease, cancer, sex, ethnicity, urbanicity, education, and the participants' age at baseline, with the exclusion of the stratified variable as appropriate. The boxes represent point estimations. Horizontal lines represent 95% CI.

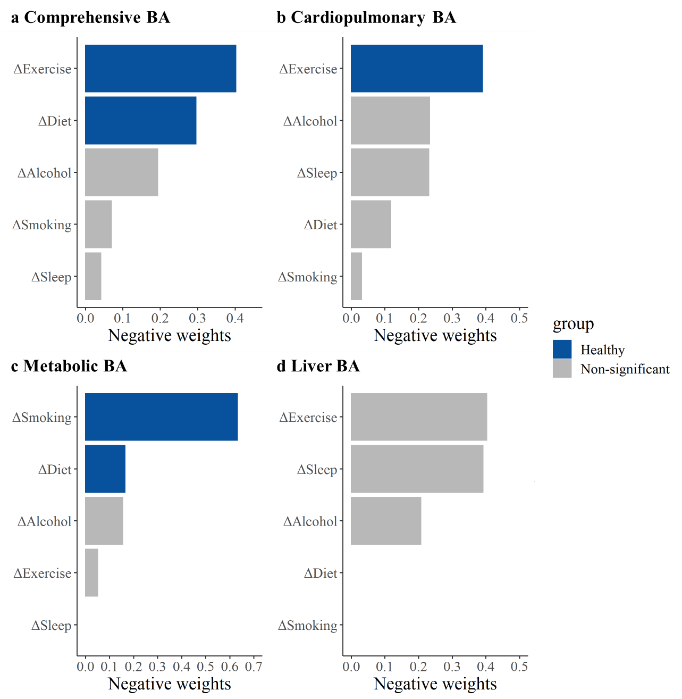

**Supplementary Figure 7. Relative contributions of five healthy lifestyle components to BAs acceleration with adjustment for time-varying covariate**

Panels: Results of the comprehensive BA acceleration, the cardiopulmonary BA acceleration, the metabolic BA acceleration, and the liver BA acceleration (A–D). ΔSmoking, change in smoking status between the baseline and repeated survey; ΔAlcohol, change in alcohol consumption between the baseline and repeated survey; ΔDiet, change in dietary quality between the baseline and repeated survey; ΔExercise, change in exercise between the baseline and repeated survey; ΔSleep, change in sleep between the baseline and repeated survey.

Estimates were obtained using QGC treating the BA accelerations as the dependent variables and five individual lifestyle factors as the independent variables. Models were adjusted for age, occupation, marital status, total energy intake, depression symptoms, anxiety symptoms, menopausal status in women, beverage intake, dietary supplement intake, diabetes, cardiovascular disease, and cancer.

The blue bars represent results that are statistically significant in the FEM analysis, while the gray bars represent results in the FEM analysis that were not found to be statistically significant and positive weights were not shown.

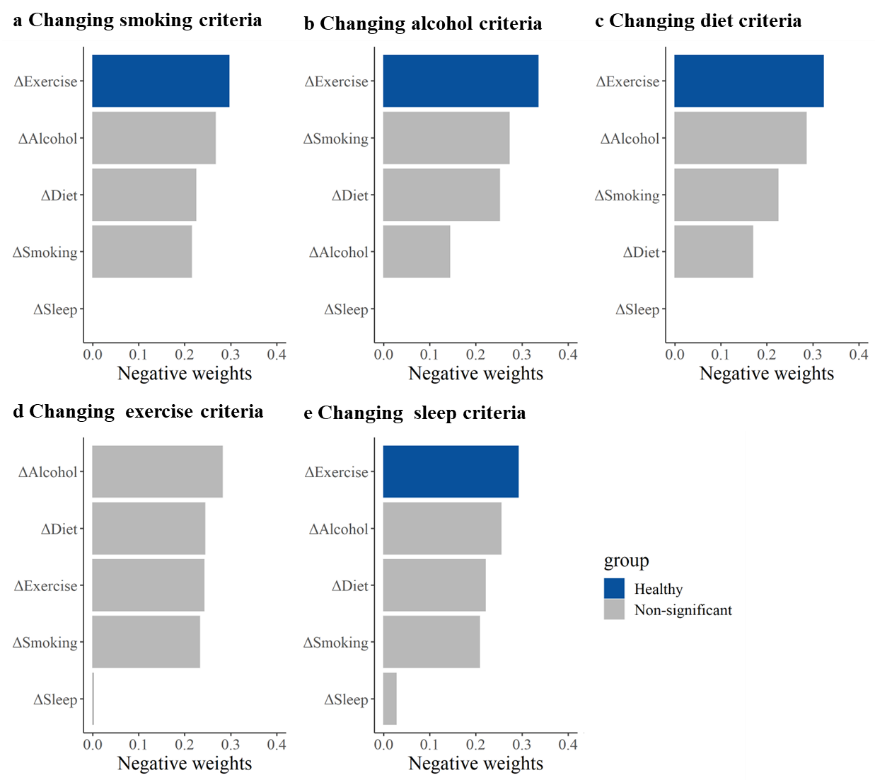

**Supplementary Figure 8. The relative contributions of five healthy lifestyle components to the comprehensive BA acceleration with altered healthy lifestyle criteria**

Panels: Results of the comprehensive BA acceleration when definitions of smoking, alcohol, diet, exercise, and sleep are changed separately (A–E). ΔSmoking, change in smoking status between the baseline and repeated survey; ΔAlcohol, change in alcohol consumption between the baseline and repeated survey; ΔDiet, change in dietary quality between the baseline and repeated survey; ΔExercise, change in exercise between the baseline and repeated survey; ΔSleep, change in sleep between the baseline and repeated survey.

Estimates were obtained using QGC treating the comprehensive BA acceleration as the dependent variables and five individual lifestyle factors as the independent variables. All models were adjusted for age, occupation, marital status, total energy intake, depression symptoms, anxiety symptoms, menopausal status in women, beverage intake, dietary supplement intake, diabetes, cardiovascular disease, cancer, sex, ethnicity, urbanicity, education, and the participants' age at baseline. The blue bars represent results that are statistically significant in the FEM analysis, while the gray bars represent results in the FEM analysis that were not found to be statistically significant and positive weights were not shown.

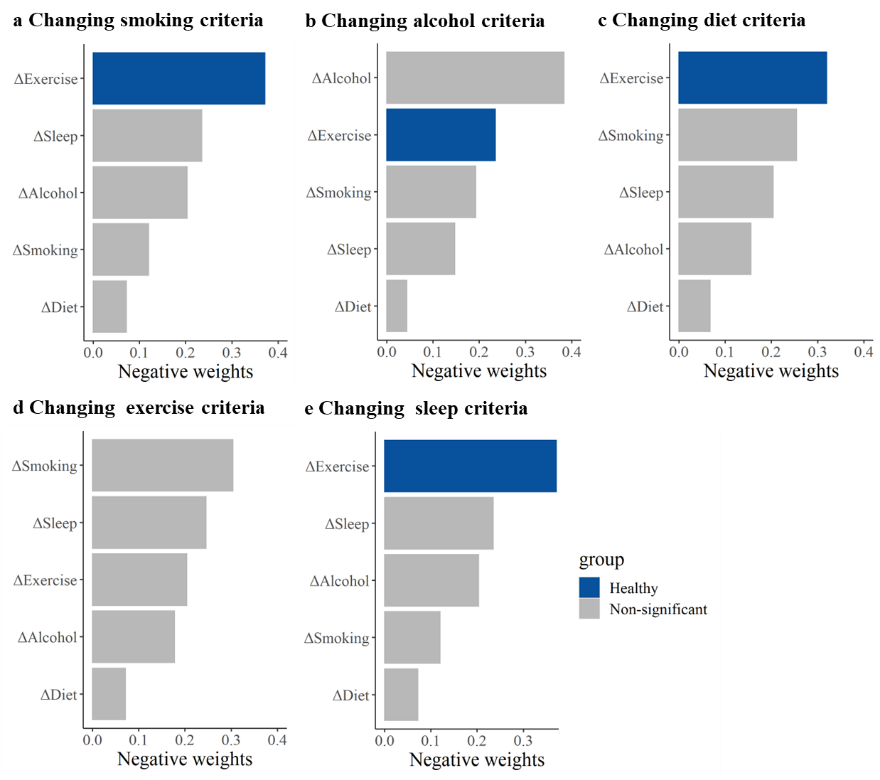

**Supplementary Figure 9. The relative contributions of five healthy lifestyle components to the cardiopulmonary BA acceleration with altered healthy lifestyle criteria**

Panels: Results of the cardiopulmonary BA acceleration when definitions of smoking, alcohol, diet, exercise, and sleep are changed separately (A–E). ΔSmoking, change in smoking status between the baseline and repeated survey; ΔAlcohol, change in alcohol consumption between the baseline and repeated survey; ΔDiet, change in dietary quality between the baseline and repeated survey; ΔExercise, change in exercise between the baseline and repeated survey; ΔSleep, change in sleep between the baseline and repeated survey.

Estimates were obtained using QGC treating the cardiopulmonary BA acceleration as the dependent variables and five individual lifestyle factors as the independent variables. All models were adjusted for age, occupation, marital status, total energy intake, depression symptoms, anxiety symptoms, menopausal status in women, beverage intake, dietary supplement intake, diabetes, cardiovascular disease, cancer, sex, ethnicity, urbanicity, education, and the participants' age at baseline. The blue bars represent results that are statistically significant in the FEM analysis, while the gray bars represent results in the FEM analysis that were not found to be statistically significant and positive weights were not shown.

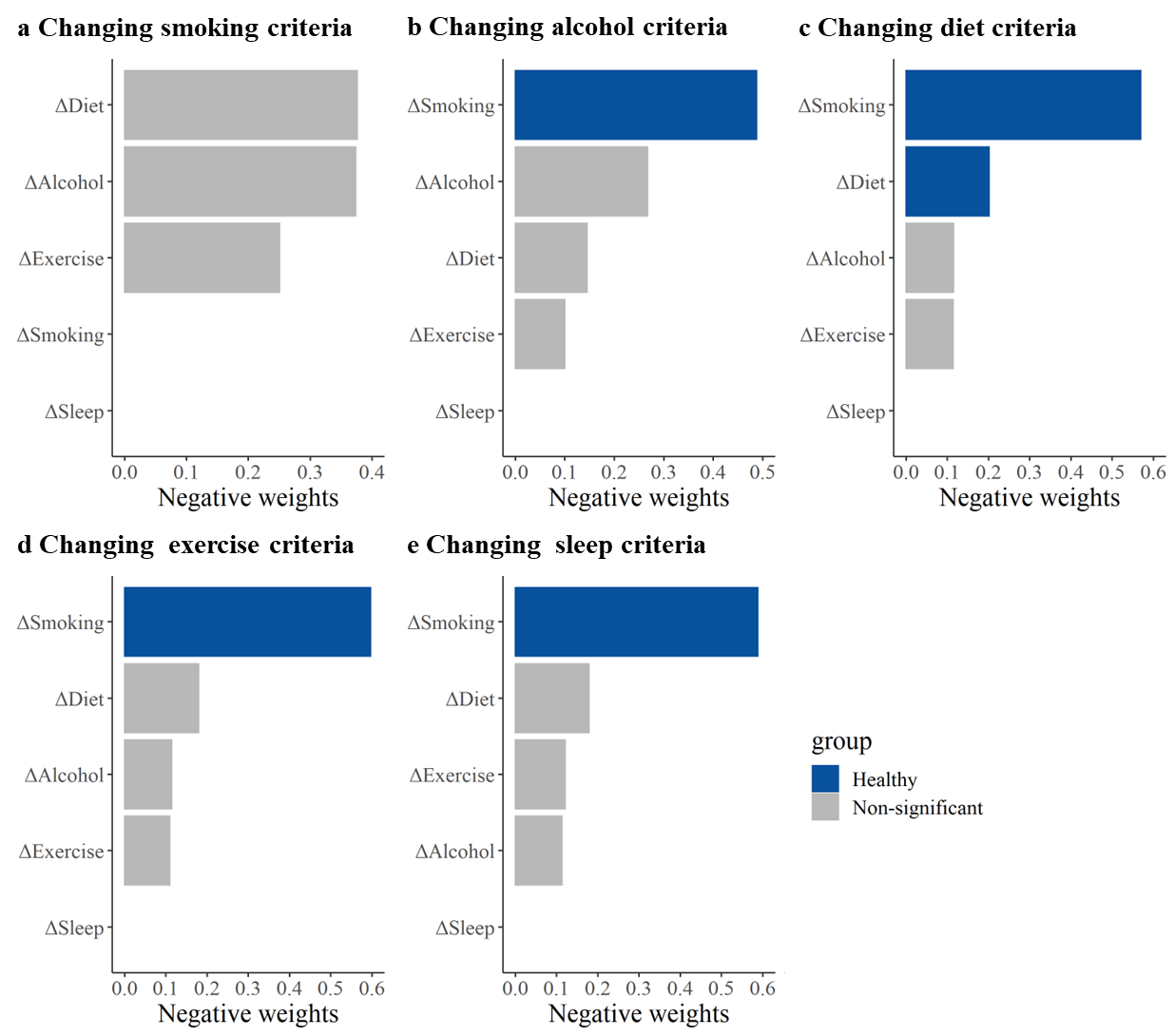

**Supplementary Figure 10. The relative contributions of five healthy lifestyle components to the metabolic BA acceleration with altered healthy lifestyle criteria**

Panels: Results of the metabolic BA acceleration when definitions of smoking, alcohol, diet, exercise, and sleep are changed separately (A–E). ΔSmoking, change in smoking status between the baseline and repeated survey; ΔAlcohol, change in alcohol consumption between the baseline and repeated survey; ΔDiet, change in dietary quality between the baseline and repeated survey; ΔExercise, change in exercise between the baseline and repeated survey; ΔSleep, change in sleep between the baseline and repeated survey.

Estimates were obtained using QGC treating the metabolic BA acceleration as the dependent variables and five individual lifestyle factors as the independent variables. All models were adjusted for age, occupation, marital status, total energy intake, depression symptoms, anxiety symptoms, menopausal status in women, beverage intake, dietary supplement intake, diabetes, cardiovascular disease, cancer, sex, ethnicity, urbanicity, education, and the participants' age at baseline. The blue bars represent results that are statistically significant in the FEM analysis, while the gray bars represent results in the FEM analysis that were not found to be statistically significant and positive weights were not shown.

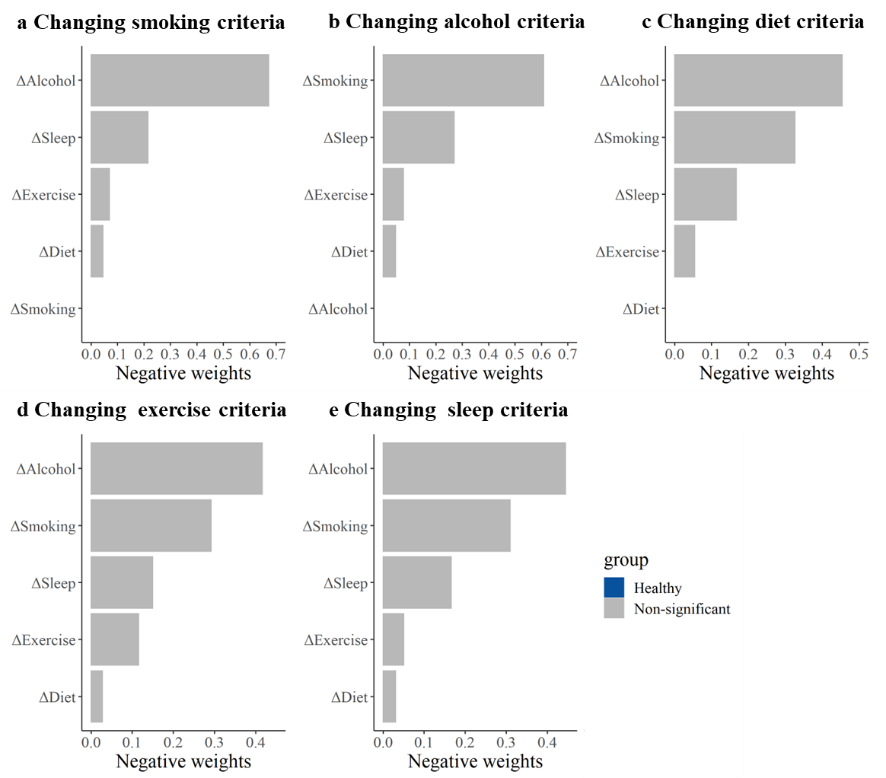

**Supplementary Figure 11. The relative contributions of five healthy lifestyle components to the liver BA acceleration with altered healthy lifestyle criteria**

Panels: Results of the liver BA acceleration when definitions of smoking, alcohol, diet, exercise, and sleep are changed separately (A–E). ΔSmoking, change in smoking status between the baseline and repeated survey; ΔAlcohol, change in alcohol consumption between the baseline and repeated survey; ΔDiet, change in dietary quality between the baseline and repeated survey; ΔExercise, change in exercise between the baseline and repeated survey; ΔSleep, change in sleep between the baseline and repeated survey.

Estimates were obtained using QGC treating the liver BA acceleration as the dependent variables and five individual lifestyle factors as the independent variables. All models were adjusted for age, occupation, marital status, total energy intake, depression symptoms, anxiety symptoms, menopausal status in women, beverage intake, dietary supplement intake, diabetes, cardiovascular disease, cancer, sex, ethnicity, urbanicity, education, and the participants' age at baseline. The blue bars represent results that are statistically significant in the FEM analysis, while the gray bars represent results in the FEM analysis that were not found to be statistically significant and positive weights were not shown.

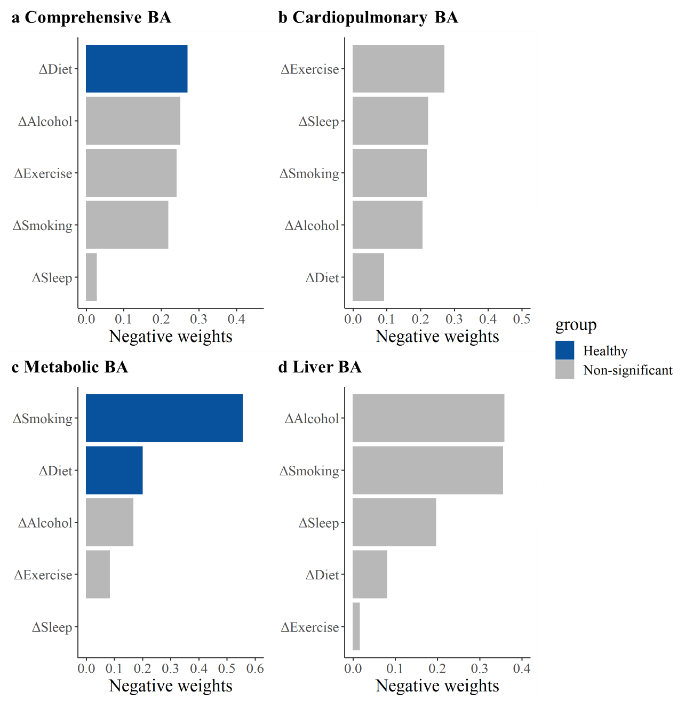

**Supplementary Figure 12. Relative contributions of five healthy lifestyle components to the comprehensive and the metabolic BA acceleration with additionally adjusted for BMI**

Panels: Results of the comprehensive BA acceleration, the cardiopulmonary BA acceleration, the metabolic BA acceleration, and the liver BA acceleration (A–D). ΔSmoking, change in smoking status between the baseline and repeated survey; ΔAlcohol, change in alcohol consumption between the baseline and repeated survey; ΔDiet, change in dietary quality between the baseline and repeated survey; ΔExercise, change in exercise between the baseline and repeated survey; ΔSleep, change in sleep between the baseline and repeated survey; BMI, body mass index.

Estimates were obtained using QGC treating the BA accelerations as the dependent variables and five individual lifestyle factors as the independent variables. Models were adjusted for age, occupation, marital status, total energy intake, depression symptoms, anxiety symptoms, menopausal status in women, beverage intake, dietary supplement intake, diabetes, cardiovascular disease, cancer, sex, ethnicity, urbanicity, education, and the participants' age at baseline.

The blue bars represent results that are statistically significant in the FEM analysis, while the gray bars represent results in the FEM analysis that were not found to be statistically significant and positive weights were not shown.

**Supplementary tables**

**Supplementary Table 1. Detailed definitions of lifestyle factors in the main analysis and sensitivity analysis**

| Exposure variable | Definitions | Definitions in sensitivity analysis |
| --- | --- | --- |
| Smoking | Never smoking was defined as healthy. | Being healthy was defined as being a non-smoker currently. |
| Alcohol consumption | Being healthy was defined as being a non-regular drinker (drinking frequency less than once a week) | Being healthy was defined as men consuming no more than 30g of pure alcohol per day and women consuming no more than 15g per day. |
| Diet | Individuals with aMED scores ≥ population median were defined as having healthy diets. | Individuals with PDI scores ≥ population median were defined as healthy diets. |
| Exercise | Based on how often participants participated in physical activity in their leisure time during the past year, regular exercise (“12 times/week”, “3–5 times/week,” or “daily or almost every day”) was categorized as healthy | Based on the time spent in each physical activity and the conventional metabolic equivalents of task (METs), MET-minutes/week were calculated. A level of ≥500 MET-minutes/week was defined as healthy. |
| Sleep | A sleep duration of 7-8 hours was defined as healthy sleep. | A sleep duration of 6-8 hours was defined as healthy sleep. |

**Supplementary Table 2. Candidate indicators used to construct the comprehensive and multi organ systems of biological age**

| **Organ/systems** | **Candidate indicators** |
| --- | --- |
| Cardiopulmonary | Systolic Blood Pressure, Diastolic Blood Pressure, Heart Rate, Peak Expiratory Flow, Total Cholesterol, High-Density Lipoprotein Cholesterol, Low-Density Lipoprotein Cholesterol, and Triglycerides. |
| Metabolism | Glycated Hemoglobin, Blood Glucose, Total Cholesterol, High-Density Lipoprotein Cholesterol, Low-Density Lipoprotein Cholesterol, Triglycerides, Body Mass Index, and Waist-Hip Ratio |
| Liver | γ-Glutamyl transpeptidase, Aspartate Aminotransferase, Alanine Aminotransferase, Alkaline Phosphatase, Total Bilirubin, Total Protein, and Albumin. |
| Renal | Creatinine, Urea, Uric Acid, and Albumin. |
| Immune systems | White Blood Cell Count, Monocyte Percentage, Monocyte Count, Lymphocyte Percentage, Lymphocyte Count, Platelet Count, Mean Platelet Volume, Platelet Distribution Width, Basophil Percentage, Basophil Count, Eosinophil Percentage, Eosinophil Count, Neutrophil Percentage, Neutrophil Count, Red Blood Cell Count, Red Cell Distribution Width, Hematocrit, Mean Corpuscular Volume, Mean Corpuscular Hemoglobin, Mean Corpuscular Hemoglobin Concentration, and Hemoglobin. |

**Supplementary Table 3 Description of BA and BA acceleration^1^**

| BA | Comprehensive  BA | Cardiopulmonary  BA | Metabolic  BA | Liver  BA | Renal BA | Immune BA |
| --- | --- | --- | --- | --- | --- | --- |
| KDM-BA |  |  |  |  |  |  |
| Baseline | 51.66 (12.45) | 51.66 (11.50) | 51.66 (13.31) | 51.66 (17.35) | 51.66 (15.24) | 51.66 (14.00) |
| Follow-up | 53.85 (12.04) | 53.43 (10.84) | 53.94 (13.01) | 56.58 (17.59) | 53.59 (14.67) | 54.24 (13.51) |
| Change | 2.49 (4.09) | 2.00 (0.94) | 2.32 (5.26) | 5.09 (12.57) | 2.31 (9.11) | 2.68 (5.07) |
| KDM-BA acceleration |  |  |  |  |  |  |
| Baseline | 0.00 (4.83) | 0.00 (0.91) | -0.00 (6.76) | -0.00 (13.02) | 0.00 (10.03) | 0.00 (8.04) |
| Follow-up | 0.39 (4.89) | -0.04 (0.94) | 0.48 (6.78) | 3.11 (13.98) | 0.13 (9.85) | 0.77 (7.95) |
| Change | 0.44 (4.09) | -0.05 (0.86) | 0.27 (5.25) | 3.05 (12.55) | 0.26 (9.14) | 0.63 (5.06) |

BA, biological age

^1^ Data are presented as mean (standard deviation).

**Supplementary Table 4. Associations of the comprehensive and multi organ systems BA acceleration with organ-specific diseases^1^**

| BA | Diseases | Participants | Events | OR (95% CI) |
| --- | --- | --- | --- | --- |
| The comprehensive BA |  |  |  |  |
| BA acceleration, continuous^2^ | CVD | 89966 | 16584 | 1.74 (1.70, 1.77) |
| BA acceleration, categorical | CVD |  |  |  |
| \|BA acceleration\|≤1^3^ |  | 15617 | 2738 | 1.00 |
| BA acceleration<-1 |  | 38297 | 4879 | 0.64 (0.61, 0.68) |
| BA acceleration>1 |  | 36052 | 8967 | 1.67 (1.59, 1.76) |
| \|BA acceleration\|≤5^4^ |  | 64458 | 11394 | 1.00 |
| BA acceleration<-5 |  | 12585 | 1207 | 0.44 (0.42, 0.48) |
| BA acceleration>5 |  | 12923 | 3983 | 2.45 (2.34, 2.57) |
| BA acceleration, continuous | Diabetes | 89966 | 4250 | 1.97 (1.91, 2.04) |
| BA acceleration, categorical | Diabetes |  |  |  |
| \|BA acceleration\|≤1 |  | 15617 | 672 | 1.00 |
| BA acceleration<-1 |  | 38297 | 1052 | 0.60 (0.55, 0.67) |
| BA acceleration>1 |  | 36052 | 2526 | 1.90 (1.73, 2.07) |
| \|BA acceleration\|≤5 |  | 64458 | 2772 | 1.00 |
| BA acceleration<-5 |  | 12585 | 240 | 0.42 (0.37, 0.48) |
| BA acceleration>5 |  | 12923 | 1238 | 3.09 (2.86, 3.33) |
| BA acceleration, continuous | Cancer | 89966 | 741 | 1.01 (0.94, 1.08) |
| BA acceleration, categorical | Cancer |  |  |  |
| \|BA acceleration\|≤1 |  | 15617 | 102 | 1.00 |
| BA acceleration<-1 |  | 38297 | 325 | 1.22 (0.98, 1.53) |
| BA acceleration>1 |  | 36052 | 314 | 1.26 (1.01, 1.58) |
| \|BA acceleration\|≤5 |  | 64458 | 506 | 1.00 |
| BA acceleration<-5 |  | 12585 | 120 | 1.09 (0.89, 1.33) |
| BA acceleration>5 |  | 12923 | 115 | 1.06 (0.86, 1.30) |
| The cardiopulmonary BA |  |  |  |  |
| BA acceleration, continuous | CVD | 89966 | 16584 | 1.46 (1.43, 1.48) |
| BA acceleration, continuous | Chronic bronchitis | 89966 | 5722 | 0.98 (0.95, 1.01) |
| The metabolic BA |  |  |  |  |
| BA acceleration, continuous | CVD | 89966 | 16584 | 1.21 (1.19, 1.23) |
| BA acceleration, categorical | CVD |  |  |  |
| \|BA acceleration\|≤1 |  | 11783 | 2047 | 1.00 |
| BA acceleration<-1 |  | 39769 | 6591 | 0.91 (0.85, 0.96) |
| BA acceleration>1 |  | 38414 | 7946 | 1.28 (1.21, 1.36) |
| \|BA acceleration\|≤5 |  | 51484 | 9139 | 1.00 |
| BA acceleration<-5 |  | 19534 | 3176 | 0.83 (0.79, 0.87) |
| BA acceleration>5 |  | 18948 | 4269 | 1.38 (1.32, 1.44) |
| BA acceleration, continuous | Diabetes | 89966 | 4250 | 3.23 (3.12, 3.34) |
| BA acceleration, categorical | Diabetes |  |  |  |
| \|BA acceleration\|≤1 |  | 11783 | 312 | 1.00 |
| BA acceleration<-1 |  | 39769 | 733 | 0.62 (0.54, 0.71) |
| BA acceleration>1 |  | 38414 | 3205 | 3.89 (3.45, 4.39) |
| \|BA acceleration\|≤5 |  | 51484 | 1479 | 1.00 |
| BA acceleration<-5 |  | 19534 | 298 | 0.44 (0.39, 0.50) |
| BA acceleration>5 |  | 18948 | 2473 | 6.33 (5.89, 6.80) |
| The liver BA |  |  |  |  |
| BA acceleration, continuous | Chronic hepatitis or cirrhosis | 89966 | 2503 | 1.28 (1.23, 1.33) |
| BA acceleration, categorical | Chronic hepatitis or cirrhosis |  |  |  |
| \|BA acceleration\|≤1 |  | 6210 | 163 | 1.00 |
| BA acceleration<-1 |  | 42219 | 946 | 0.80 (0.68, 0.95) |
| BA acceleration>1 |  | 41537 | 1394 | 1.20 (1.02, 1.42) |
| \|BA acceleration\|≤5 |  | 29686 | 728 | 1.00 |
| BA acceleration<-5 |  | 30423 | 671 | 0.83 (0.75, 0.93) |
| BA acceleration>5 |  | 29857 | 1104 | 1.40 (1.27, 1.55) |
| The immune BA |  |  |  |  |
| BA acceleration, continuous | Rheumatoid arthritis | 89966 | 2545 | 0.97 (0.93, 1.00) |
| BA acceleration, categorical | Rheumatoid arthritis |  |  |  |
| \|BA acceleration\|≤1 |  | 11083 | 281 | 1.00 |
| BA acceleration<-1 |  | 38363 | 1211 | 1.10 (0.97, 1.26) |
| BA acceleration>1 |  | 40520 | 1053 | 1.07 (0.93, 1.22) |
| \|BA acceleration\|≤5 |  | 49112 | 1354 | 1.00 |
| BA acceleration<-5 |  | 19355 | 667 | 1.07 (0.97, 1.18) |
| BA acceleration>5 |  | 21499 | 524 | 0.93 (0.84, 1.04) |

Abbreviation: CVD, cardiovascular disease.

^1^ Participants with available data on baseline BA, self-report diseases, and covariates (n=89966) were included for validation analysis. Logistic regression models were used. Results were adjusted for five individual healthy lifestyle factors (smoking, alcohol consumption, diet, exercise, and sleep), as well as for age, sex, ethnicity, urbanicity, education, occupation, marital status, total energy intake, symptoms of depression and anxiety, menopausal status in women, beverage intake, dietary supplement intake, and body mass index (BMI) as covariates. For the cardiopulmonary BA, due to the small values of BA acceleration, no categorized analysis was conducted. Since the values of BA acceleration of the cardiopulmonary BA were small, it was not processed as categorical variables.

^2^ Per 1-SD increase (or improvement) in BA acceleration

^3^ Represents the reference group

^4^ Represents the reference group

**Supplementary Table 5. Associations of healthy lifestyle factors and HLI with the BA accelerations with adjustment for time-varying covariates**

| **Variables** | **Comprehensive**  **BA acceleration** | **Cardiopulmonary**  **BA acceleration** | **Metabolic**  **BA acceleration** | **Liver**  **BA acceleration** |
| --- | --- | --- | --- | --- |
|  | $\boldsymbol{\beta}$ **(95%CI)** | $\boldsymbol{\beta}$ **(95%CI)** | $\boldsymbol{\beta}$ **(95%CI)** | $\boldsymbol{\beta}$ **(95%CI)** |
| HLI (range) |  |  |  |  |
| Per 1-point increase | -0.11 (-0.19, -0.03) | -0.02 (-0.04, -0.00) | -0.15 (-0.25, -0.05) | -0.13 (-0.37, 0.11) |
| HLI (category) |  |  |  |  |
| To have a healthy lifestyle | -0.18 (-0.34, -0.03) | -0.03 (-0.06, 0.01) | -0.19 (-0.39, 0.01) | -0.11 (-0.59, 0.37) |
| Healthy lifestyle factors |  |  |  |  |
| ΔSmoking | -0.04 (-0.37, 0.30) | -0.00 (-0.07, 0.07) | -0.86 (-1.29, -0.42) | 0.94 (-0.09, 1.98) |
| ΔAlcohol | -0.10 (-0.40, 0.21) | -0.02 (-0.09, 0.04) | -0.21 (-0.60, 0.18) | -0.17 (-1.11, 0.77) |
| ΔDiet | -0.15 (-0.30, -0.01) | -0.01 (-0.04, 0.02) | -0.22 (-0.41, -0.04) | 0.03 (-0.41, 0.48) |
| ΔExercise | -0.20 (-0.37, -0.04) | -0.04 (-0.07, -0.00) | -0.07 (-0.28, 0.14) | -0.33 (-0.83, 0.17) |
| ΔSleep | -0.02 (-0.16, 0.12) | -0.02 (-0.05, 0.01) | 0.00 (-0.18, 0.19) | -0.32 (-0.76, 0.11) |

BA, biological age; HLI, healthy lifestyle indicator.

ΔSmoking, change in smoking status between the baseline and repeated survey; ΔAlcohol, change in alcohol consumption between the baseline and repeated survey; ΔDiet, change in dietary quality between the baseline and repeated survey; ΔExercise, change in exercise between the baseline and repeated survey; ΔSleep, change in sleep between the baseline and repeated survey; CI, confidence interval.

Estimates were obtained using FEMs treating the BA accelerations as the dependent variables and HLI (as either continuous or as categorized) or five individual lifestyle factors as the independent variables. Models were adjusted for age, occupation, marital status, total energy intake, depression symptoms, anxiety symptoms, menopausal status in women, beverage intake, dietary supplement intake, diabetes, cardiovascular disease, cancer.

**Supplementary Table 6. Associations of healthy lifestyle factors and HLI with the BA accelerations altered healthy lifestyle criteria**

| Variables | Changing smoking criteria | Changing alcohol criteria | Changing diet  criteria | Changing exercise criteria | Changing sleep criteria |
| --- | --- | --- | --- | --- | --- |
|  | β (95%CI) | β (95%CI) | β (95%CI) | β (95%CI) | β (95%CI) |
| The comprehensive BA acceleration |  |  |  |  |  |
| HLI (range) |  |  |  |  |  |
| Per 1-point increase | -0.10 (-0.18, -0.02) | -0.10 (-0.18, -0.02) | -0.09 (-0.17, -0.01) | -0.09 (-0.17, -0.01) | -0.11 (-0.19, -0.03) |
| HLI (category) |  |  |  |  |  |
| To have a healthy lifestyle | -0.19 (-0.35, -0.03) | -0.15 (-0.31, 0.00) | -0.09 (-0.24, 0.06) | -0.16 (-0.32, -0.00) | -0.19 (-0.34, -0.03) |
| Healthy lifestyle factors |  |  |  |  |  |
| ΔSmoking | -0.13 (-0.55, 0.29) | -0.15 (-0.49, 0.20) | -0.13 (-0.47, 0.22) | -0.13 (-0.48, 0.22) | -0.13 (-0.48, 0.22) |
| ΔAlcohol | -0.16 (-0.47, 0.15) | -0.08 (-0.51, 0.35) | -0.16 (-0.47, 0.15) | -0.16 (-0.47, 0.15) | -0.16 (-0.47, 0.15) |
| ΔDiet | -0.14 (-0.28, 0.01) | -0.14 (-0.28, 0.01) | -0.09 (-0.24, 0.05) | -0.14 (-0.28, 0.01) | -0.14 (-0.28, 0.01) |
| ΔExercise | -0.18 (-0.35, -0.02) | -0.18 (-0.35, -0.02) | -0.18 (-0.35, -0.01) | -0.14 (-0.30, 0.03) | -0.18 (-0.35, -0.02) |
| ΔSleep | 0.00 (-0.14, 0.15) | 0.00 (-0.14, 0.14) | 0.00 (-0.14, 0.15) | -0.00 (-0.14, 0.14) | -0.02 (-0.16, 0.13) |
| The cardiopulmonary BA acceleration |  |  |  |  |  |
| HLI (range) |  |  |  |  |  |
| Per 1-point increase | -0.02 (-0.04, -0.00) | -0.02 (-0.04, -0.01) | -0.02 (-0.04, -0.01) | -0.02 (-0.04, -0.00) | -0.02 (-0.04, -0.01) |
| HLI (category) |  |  |  |  |  |
| To have a healthy lifestyle | -0.03 (-0.06, 0.00) | -0.03 (-0.06, 0.00) | -0.03 (-0.06, 0.00) | -0.02 (-0.06, 0.01) | -0.03 (-0.07, -0.00) |
| Healthy lifestyle factors |  |  |  |  |  |
| ΔSmoking | -0.01 (-0.10, 0.08) | -0.03 (-0.10, 0.04) | -0.03 (-0.10, 0.04) | -0.03 (-0.10, 0.04) | -0.03 (-0.10, 0.04) |
| ΔAlcohol | -0.02 (-0.09, 0.04) | -0.06 (-0.15, 0.03) | -0.02 (-0.08, 0.05) | -0.02 (-0.08, 0.05) | -0.02 (-0.08, 0.05) |
| ΔDiet | -0.01 (-0.04, 0.02) | -0.01 (-0.04, 0.02) | -0.01 (-0.04, 0.02) | -0.01 (-0.04, 0.02) | -0.01 (-0.04, 0.02) |
| ΔExercise | -0.04 (-0.07, -0.00) | -0.04 (-0.07, -0.00) | -0.04 (-0.07, -0.00) | -0.02 (-0.06, 0.01) | -0.04 (-0.07, -0.00) |
| ΔSleep | -0.02 (-0.05, 0.01) | -0.02 (-0.05, 0.01) | -0.02 (-0.05, 0.01) | -0.03 (-0.06, 0.01) | -0.03 (-0.06, 0.00) |
| The metabolic BA acceleration |  |  |  |  |  |
| HLI (range) |  |  |  |  |  |
| Per 1-point increase | -0.07 (-0.18, 0.03) | -0.11 (-0.21, -0.00) | -0.11 (-0.21, -0.01) | -0.10 (-0.20, 0.00) | -0.10 (-0.21, -0.00) |
| HLI (category) |  |  |  |  |  |
| To have a healthy lifestyle | -0.14 (-0.34, 0.07) | -0.10 (-0.30, 0.10) | -0.07 (-0.26, 0.12) | -0.15 (-0.35, 0.06) | -0.13 (-0.33, 0.08) |
| Healthy lifestyle factors |  |  |  |  |  |
| ΔSmoking | 0.02 (-0.52, 0.55) | -0.55 (-0.99, -0.11) | -0.53 (-0.97, -0.09) | -0.54 (-0.98, -0.10) | -0.54 (-0.98, -0.10) |
| ΔAlcohol | -0.17 (-0.56, 0.23) | -0.30 (-0.85, 0.25) | -0.11 (-0.50, 0.29) | -0.10 (-0.50, 0.29) | -0.10 (-0.50, 0.29) |
| ΔDiet | -0.17 (-0.35, 0.02) | -0.16 (-0.35, 0.02) | -0.19 (-0.38, -0.00) | -0.16 (-0.35, 0.02) | -0.16 (-0.35, 0.02) |
| ΔExercise | -0.11 (-0.32, 0.10) | -0.11 (-0.32, 0.10) | -0.11 (-0.32, 0.10) | -0.10 (-0.31, 0.11) | -0.11 (-0.32, 0.10) |
| ΔSleep | 0.05 (-0.14, 0.23) | 0.04 (-0.14, 0.22) | 0.04 (-0.14, 0.22) | 0.04 (-0.14, 0.22) | 0.04 (-0.14, 0.22) |
| The liver BA acceleration |  |  |  |  |  |
| HLI (range) | -0.19 (-0.43, 0.05) | -0.16 (-0.41, 0.08) | -0.15 (-0.39, 0.09) | -0.26 (-0.50, -0.02) | -0.23 (-0.47, 0.01) |
| Per 1-point increase | -0.18 (-0.65, 0.29) | -0.09 (-0.56, 0.38) | -0.18 (-0.63, 0.27) | -0.25 (-0.73, 0.22) | -0.21 (-0.68, 0.27) |
| HLI (category) |  |  |  |  |  |
| To have a healthy lifestyle |  |  |  |  |  |
| Healthy lifestyle factors |  |  |  |  |  |
| ΔSmoking | 0.19 (-1.06, 1.43) | -0.67 (-1.70, 0.35) | -0.57 (-1.60, 0.46) | -0.56 (-1.59, 0.47) | -0.56 (-1.59, 0.47) |
| ΔAlcohol | -0.88 (-1.80, 0.04) | 0.31 (-0.97, 1.59) | -0.79 (-1.72, 0.13) | -0.80 (-1.72, 0.12) | -0.80 (-1.72, 0.12) |
| ΔDiet | -0.06 (-0.50, 0.38) | -0.05 (-0.49, 0.39) | 0.19 (-0.25, 0.62) | -0.05 (-0.49, 0.39) | -0.05 (-0.49, 0.38) |
| ΔExercise | -0.09 (-0.58, 0.40) | -0.08 (-0.58, 0.41) | -0.09 (-0.59, 0.40) | -0.22 (-0.72, 0.28) | -0.09 (-0.58, 0.40) |
| ΔSleep | -0.28 (-0.71, 0.15) | -0.30 (-0.73, 0.13) | -0.29 (-0.72, 0.14) | -0.29 (-0.72, 0.14) | -0.30 (-0.73, 0.13) |

BA, biological age; HLI, healthy lifestyle indicator.

ΔSmoking, change in smoking status between the baseline and repeated survey; ΔAlcohol, change in alcohol consumption between the baseline and repeated survey; ΔDiet, change in dietary quality between the baseline and repeated survey; ΔExercise, change in exercise between the baseline and repeated survey; ΔSleep, change in sleep between the baseline and repeated survey; CI, confidence interval.

Estimates were obtained using FEMs treating the BA accelerations as the dependent variables and HLI (as either continuous or as categorized) or five lifestyle factors as the independent variables. All models were adjusted for age, occupation, marital status, total energy intake, depression symptoms, anxiety symptoms, menopausal status in women, beverage intake, dietary supplement intake, diabetes, cardiovascular disease, cancer, sex, ethnicity, urbanicity, education, and the participants' age at baseline.

**Supplementary Table 7. Associations of healthy lifestyle factors and HLI with the BA accelerations with additionally adjusted for BMI**

| Variables | Comprehensive  BA acceleration | Cardiopulmonary  BA acceleration | Metabolic  BA acceleration | Liver  BA acceleration |
| --- | --- | --- | --- | --- |
|  | $\beta$ (95%CI) | $\beta$ (95%CI) | $\beta$ (95%CI) | $\beta$ (95%CI) |
| HLI (range) |  |  |  |  |
| Per 1-point increase | -0.11 (-0.19, -0.03) | -0.02 (-0.04, -0.01) | -0.12 (-0.22, -0.02) | -0.22 (-0.46, 0.01) |
| HLI (category) |  |  |  |  |
| To have a healthy lifestyle | -0.18 (-0.33, -0.02) | -0.03 (-0.07, 0.00) | -0.16 (-0.36, 0.04) | -0.19 (-0.65, 0.28) |
| Healthy lifestyle factors |  |  |  |  |
| ΔSmoking | -0.13 (-0.47, 0.21) | -0.03 (-0.10, 0.05) | -0.53 (-0.96, -0.10) | -0.58 (-1.58, 0.43) |
| ΔAlcohol | -0.15 (-0.45, 0.15) | -0.02 (-0.09, 0.04) | -0.16 (-0.55, 0.23) | -0.58 (-1.48, 0.32) |
| ΔDiet | -0.16 (-0.30, -0.02) | -0.01 (-0.04, 0.02) | -0.19 (-0.37, -0.01) | -0.13 (-0.56, 0.30) |
| ΔExercise | -0.14 (-0.30, 0.02) | -0.03 (-0.07, 0.00) | -0.08 (-0.29, 0.13) | -0.02 (-0.51, 0.46) |
| ΔSleep | -0.02 (-0.16, 0.12) | -0.03 (-0.06, 0.00) | 0.01 (-0.17, 0.19) | -0.32 (-0.74, 0.10) |

BA, biological age; HLI, healthy lifestyle indicator.

ΔSmoking, change in smoking status between the baseline and repeated survey; ΔAlcohol, change in alcohol consumption between the baseline and repeated survey; ΔDiet, change in dietary quality between the baseline and repeated survey; ΔExercise, change in exercise between the baseline and repeated survey; ΔSleep, change in sleep between the baseline and repeated survey; CI, confidence interval.

Estimates were obtained using FEMs treating the BA accelerations as the dependent variables and HLI (as either continuous or as categorized) or five individual lifestyle factors as the independent variables. Models were adjusted for age, occupation, marital status, total energy intake, depression symptoms, anxiety symptoms, menopausal status in women, beverage intake, dietary supplement intake, diabetes, cardiovascular disease, cancer, sex, ethnicity, urbanicity, education, and the participants' age at baseline.
